# The Epithelial and Stromal Immune Microenvironment in Gastric Cancer: A Comprehensive Analysis Reveals Prognostic Factors with Digital Cytometry

**DOI:** 10.1101/2021.08.10.21261822

**Authors:** Wenjun Shen, Guoyun Wang, Georgia R. Cooper, Yuming Jiang, Xin Zhou

**Affiliations:** Department of Bioinformatics, Shantou University Medical College, Shantou, Guangdong, China; Stanford Center for Biomedical Informatics Research (BMIR), Department of Medicine, Stanford University, Stanford, CA, USA; Guangdong Provincial Key Laboratory of Infectious Diseases and Molecular Immunopathology, Shantou, Guangdong, China; Department of Biomedical Engineering, Vanderbilt University, Nashville, TN 37235, USA; Department of Radiation Oncology, Stanford University School of Medicine, Stanford, CA, USA; Department of Computer Science, Vanderbilt University, Nashville, TN 37235, USA

## Abstract

**Background:** Gastric cancer (GC) is the third leading cause of cancer-related deaths worldwide. Tumor heterogeneity continues to confound researchers’ understanding of tumor growth and the development of an effective therapy. Digital cytometry allows interpretation of heterogeneous bulk tissue transcriptomes at the cellular level.

**Methods:** We built a novel signature matrix to dissect epithelium and stroma signals using a scRNA-seq data set for GC. We applied cell mixture deconvolution to estimate diverse epithelial, stromal, and immune cell proportions from bulk transcriptome data in four independent GC cohorts. Robust computational methods were applied to identify strong prognostic factors for GC.

**Results:** We identified an EMEC population whose proportions were significantly higher in patients with stage I cancer than other stages, and it was predominantly present in tumor samples but not typically found in normal samples. We found that the ratio of EMECs to stromal cells and the ratio of adaptive T cells to monocytes were the most significant prognostic factors within the non-immune and immune factors, respectively. The STEM score, which unifies these two prognostic factors was an independent prognostic factor of overall survival (HR=0.92, 95% CI=0.89-0.94, *p* = 2.05 × 10^−9^). The entire GC cohort was stratified into three risk groups (high-, moderate-, and low-risk) which yielded incremental survival times (*p* < 0.0001). For stage III disease, patients in the moderate- and low-risk groups experienced better survival benefits from radiation therapy ((HR=0.16, 95% CI=0.06-0.4, *p* < 0.0001), whereas those in the high-risk group did not (HR=0.49, 95% CI=0.14-1.72, *p* = 0.25).

**Conclusions:** We conclude that the STEM score is a promising prognostic factor for gastric cancer.

## 1 Introduction

Gastric cancer (GC) is a complex and heterogeneous disease from morphological, molecular, and cellular standpoints [1]. Such tumor heterogeneity has been demonstrated in numerous histological and molecular classifications. The Lauren classification separates gastric adenocarcinomas into intestinal, diffuse, and mixed subtypes which were found to be associated with varying stomach cancer risks [2, 3]. The Asian Cancer Research Group (ACRG) classified GC into four molecular subtypes that, based on gene expression data, were associated with distinct molecular alterations, disease progression, and survival outcomes [4]. These subtypes are epithelial-to-mesenchymal transition (EMT), MSS/TP53-, MSS/TP53+, and microsatellite instability (MSI). More recently, The Cancer Genome Atlas (TCGA) research network characterized GC into four genomic subtypes by integrating data from six molecular platforms: array-based somatic copy number analysis, whole-exome sequencing, array-based DNA methylation profiling, messenger RNA sequencing, microRNA (miRNA) sequencing and reverse-phase protein array, as well as Microsatellite instability (MSI) testing [5]. These genomic subtypes are EBV-positivity (EBV), MSI-high status (MSI), genomically stable (GS), and those exhibiting chromosomal instability (CIN). Each subtype displays distinct molecular and genomic patterns.

Tumor heterogeneity, including the results from the tumor microenvironment (TME), continues to confound researchers’ understanding of tumor growth and the development of an effective therapy [6,7]. Tumors are complex ecosystems that are affected by numerous stromal and immune factors which dampen or enhance the effects of genetic epithelial alterations [8–11]. The TME is comprised of tumor cells, tumor stromal cells, endothelial cells, immune cells, and the non-cellular components of extracellular matrix proteins [12, 13]. Some essential components of the TME, including cancer-associated fibroblasts (CAF) [14, 15], tumor-infiltrating lymphocytes (TIL) [16, 17], tumor-associated macrophages (TAM) [18], and other cellular components [19–21], have been evaluated to help researchers better understand the role of the TME in gastric cancer risk. Most of these studies have focused on the subsets of cellular components of TME: typically the stromal and immune cell populations. However, the prognostic values of diverse gastric epithelial cell types in GC risk were still unclear. To our knowledge, a systematic analysis of the prognostic value of diverse epithelial cell types, including cancer cell, MSC, PMC and PC, emerged in early gastric cancer for predicting survival has not been described.

Digital cytometry enables the examination of heterogenous bulk tissue transcriptomes at the cellular level in addition to using computational methods to quantify cell type composition. In recent years, single cell RNA-sequencing (scRNA-seq) techniques have offered new insights into tissue samples at the resolution of single cells. The availability of single cell transcriptomic profiles, which are used to build cell type-specific signature matrices, promote the development of statistical deconvolution methods for estimating cell type compositions in heterogeneous mixture samples. [22, 23].

This study attempts to investigate the TME of GC with comprehensive epithelial, stromal, and immune cell profiling by combining single cell and bulk expression profiles. We have undertaken a comprehensive analysis on 10 non-immune and 7 immune cell populations of the TME of GC, evaluated the prognostic role of the STEM score in four independent GC cohorts, and stratified the GC patients into three TME subtypes based on the abundance of four STEM pupulations.

## 2 Results

### 2.1 Building a non-immune signature matrix for GC from a single-cell RNA-seq data set

We carried out a systematic cell subtype analysis on a single-cell RNA-seq data set of patients with gastric premalignant lesions and early gastric cancer [24]. An unsupervised hierarchical clustering analysis was used to investigate the possibility of identifying different non-immune cell populations in the gastric scRNA-seq data set based on their expression profiles. Hierarchical clustering of the cell types in the cascade stages from non-atrophic gastritis (NAG), chronic atrophic gastritis (CAG), intestinal metaplasia (IM) and early gastric cancer (EGC) in the single cell reference revealed that 24,874 non-immune cells fell into three large groups: epithelial cell types (including cancer cell, enterocyte, enteroendocrine, GMC, goblet cell, MSC, PC, and PMC), stromal cell types (including Fibroblast and SM cells), and endothelial cell types (Fig.1). Notably, the enterocytes and cancer cells that emerged in the CAG and IM biopsies were clustered together. We labeled this cluster as the premalignant epithelial cell (PMEC) pupulation. We also detected a cell population in which cells were clustered by stage instead of by cell type. This cell population consists of four gastric epithelial cell types (including cancer cell, MSC, PC, and PMC) that emerged uniformly in the EGC biopsy. Therefore, we called it the early malignant epithelial cell (EMEC) subtype. The other eight cell populations were found to clustered by cell type. Overall, ten cell populations of EMEC, PMEC, enteroendocrine, GMC, goblet cell, MSC, PC, PMC, endothelial cell, and stromal cell were detected in the cluster tree (Fig.1, Table 5). To estimate the proportions of non-immune cell compositions in the gastric cancer samples, we created a cell population-specific signature matrix to distinguish these 10 cell populations. The cell population-specific marker genes were selected by using two-way ANOVA (see Methods for details).

**Figure 1:**
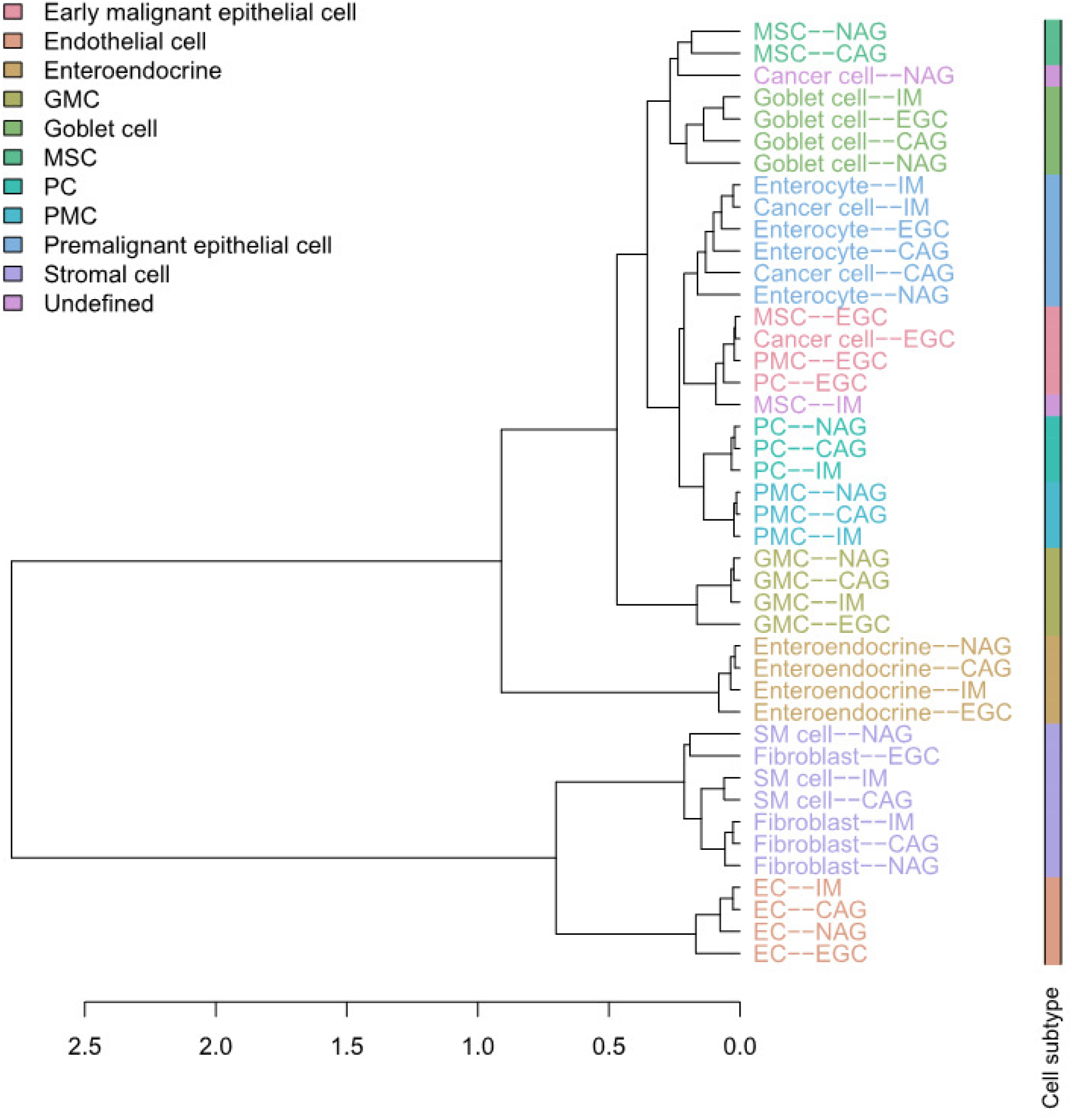
Cluster tree of 11 non-immune cell types of patients with NAG, CAG, IM and EGC.

### 2.2 Dissecting epithelium-stroma-immune signals from gastric cancer samples

Epithelial, stromal, and immune cell populations comprise the vast majority of gastric tumor cellularity. In order to accurately dissect epithelium-stroma-immune signals from GC samples, we applied CIBERSORT [25] or CIBERSORTx [23] to RNA profiles of GC samples. Our methodology involved two separate deconvolution procedures: an immune cell deconvolution procedure and a non-immune cell deconvolution procedure, which dissects immune and non-immune signals from GC samples, respectively. The immune system is an important determinant of the TME, we applied CIBERSORT/CIBERSORTx to infer relative immune cell subtype fractions in four cohorts of bulk GC samples by using a signature matrix derived from the peripheral blood mononuclear cell (PBMC) samples (Fig.2, see Methods for details). We observed that the T adaptive cell population, comprised of naive and memory CD4 and CD8 T cells, was consistently dominant across the four GC cohorts, followed by the monocyte, B adaptive, and T innate subsets. The single cell gene expression data sets of GC offer new insights to investigating the GC TME at the resolution of single cells. To enumerate GC non-immune cell proportions, we further applied CIBERSORT/CIBERSORTx by using the single-cell reference profiles to distinguish epithelial, stromal, and endothelial cell subsets in the bulk GC samples (Fig.3, see Methods for details). We observed that the EMEC, PC, and stromal cell subsets were highly abundant while the endothelial cell subset was less abundant in all four cohorts.

**Figure 2:**
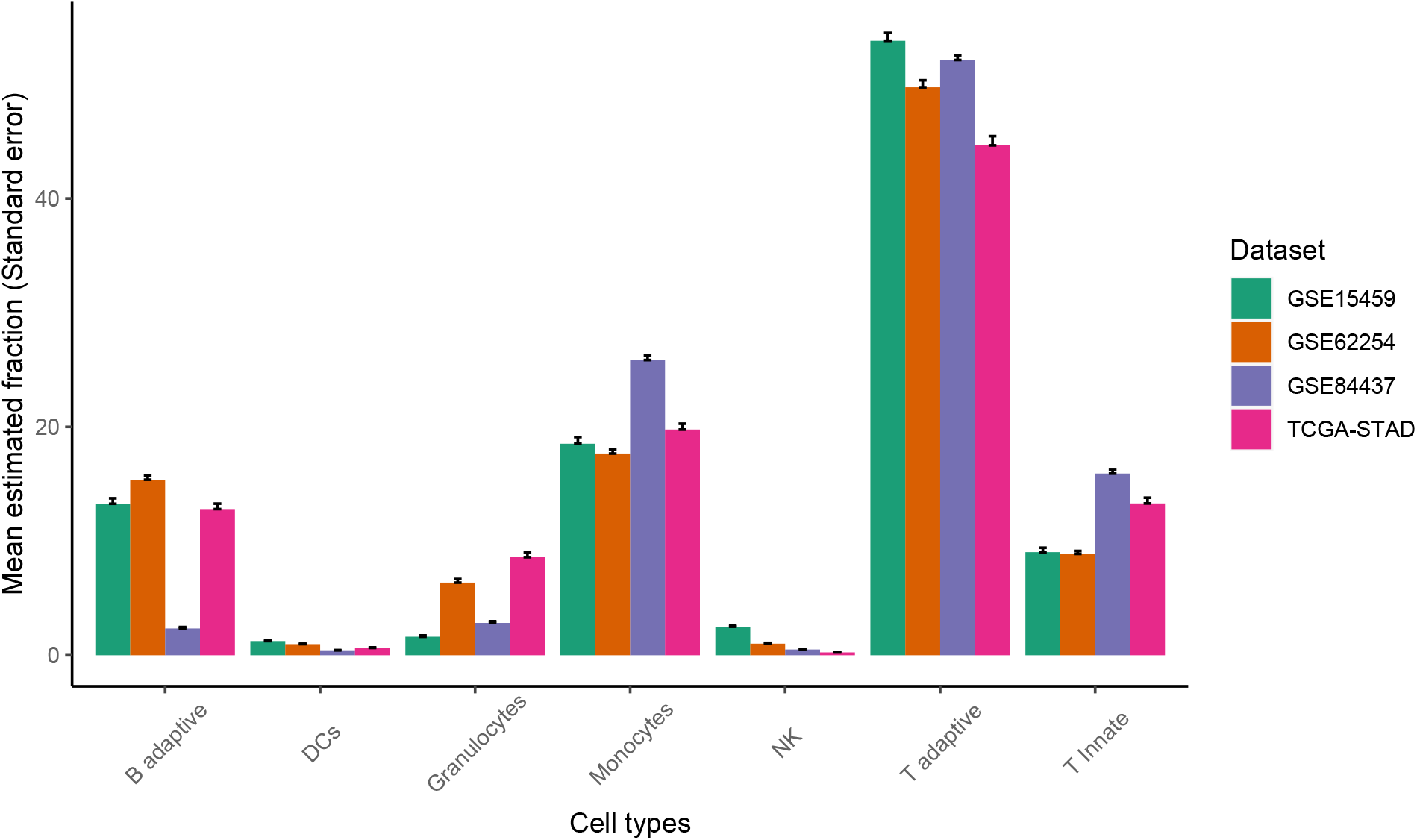
The relative abundance of major classes of immune cell populations for four cohorts of gastric cancer samples.

**Figure 3:**
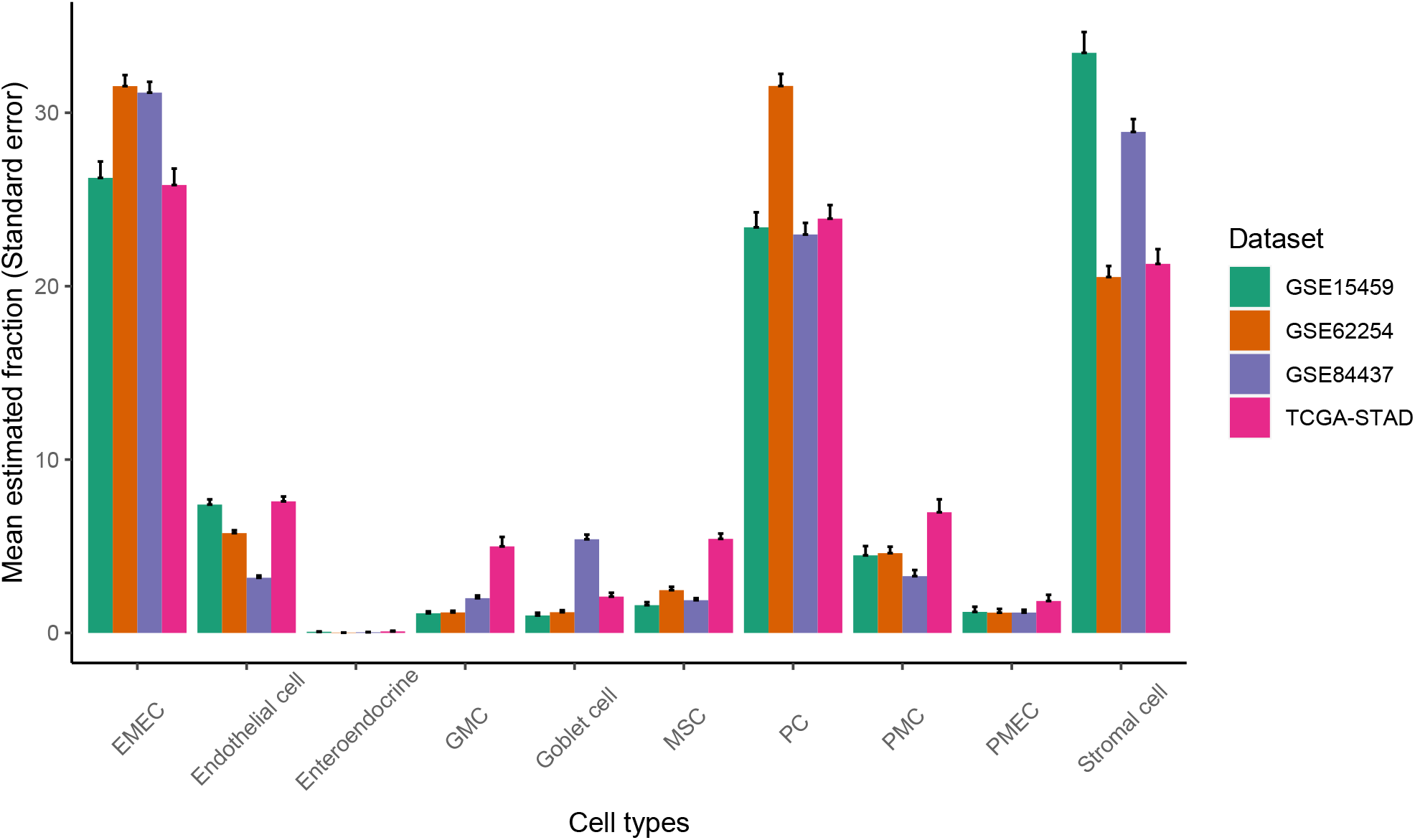
The relative abundance of major classes of non-immune cell populations for four cohorts of gastric cancer samples.

The EMEC population is composed of gastric epithelial cells (including cancer cells, MSCs, PCs, and PMCs) that were isolated from early GC patients. We analyzed the functional annotation of the EMEC gene signatures using Gene Ontology (GO). The EMEC gene signatures were mostly enriched for mitochondrial translation, mitochondrial translational elongation and mitochondrial translational termination in the biological process (BP) ontology (Fig. 4a); and were mainly enriched in the mitochondrial inner membrane and mitochondrial ribosome in the cellular component (CC) ontology (Fig. 4b). The results suggest that the mitochondria could serve as a GC biomarker for early detection, which are consistent with previous reports showing a role for mitochondria [26–28] in the early detection of solid tumors.

**Figure 4:**
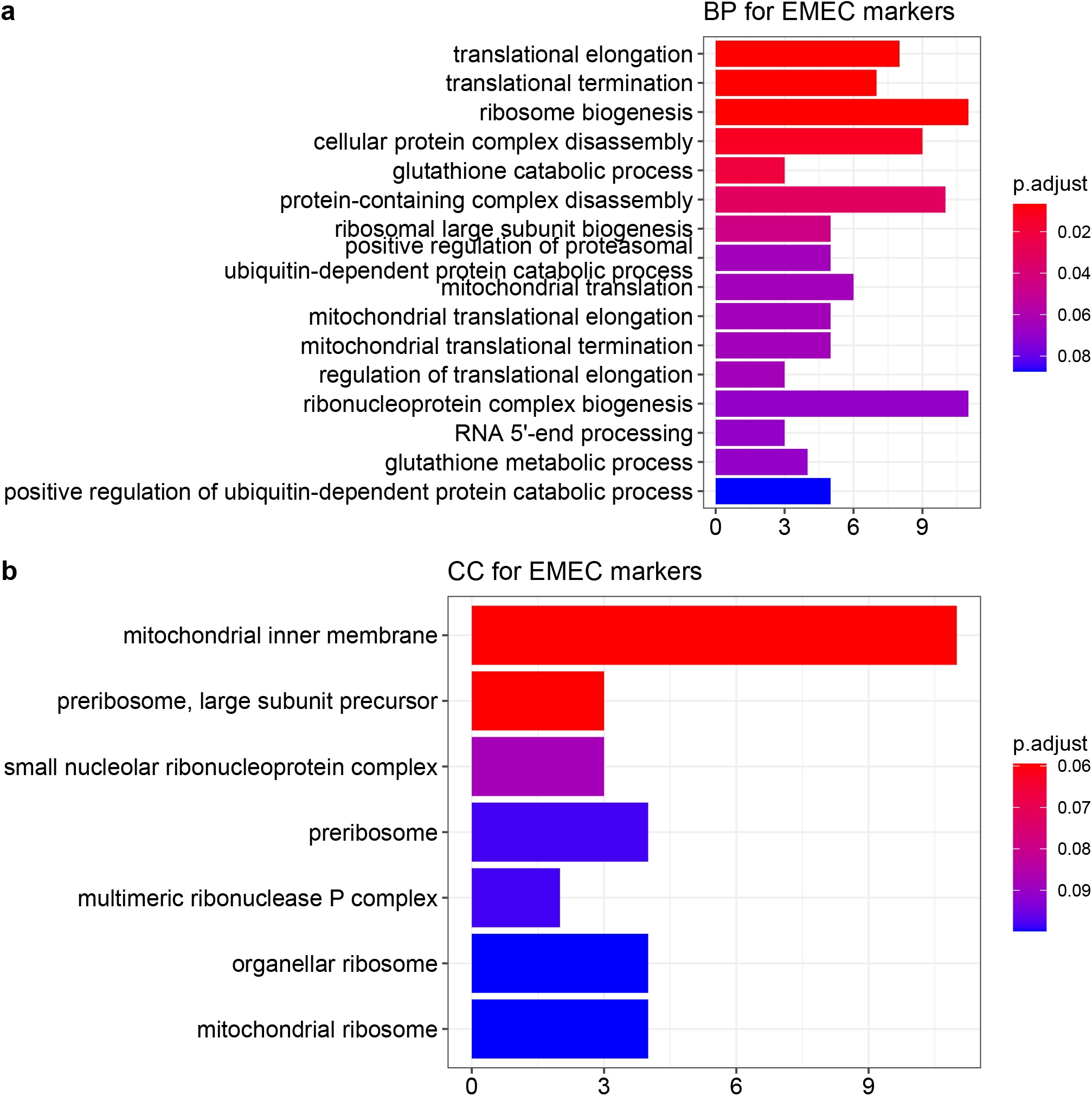
Top GO terms enriched for the EMEC gene signatures in the BP (a) and CC (b) ontologies.

We hypothesized that the EMEC population would be more abundant in patients with stage I cancer than other stages. To test this hypothesis, we estimated proportions of the EMEC population in each sample of both the ACRG and TCGA-STAD cohorts. We observed significantly higher EMEC populations in patients with stage I cancer than stage II, III, or IV in both the ACRG and TCGA-STAD cohorts (*p* < 0.05, Student’s t-test; Fig.5). To investigate whether the EMEC population was typically found in tumor samples but not in normal samples, we estimated proportions of 10 non-immune cell populations in samples from both tumor and adjacent normal tissues available in the TCGA-STAD sample collection. We observed that the EMEC population was significantly higher in tumor samples than in adjacent normal samples (*p* < 0.0001, Student’s t-test; Fig.6).

**Figure 5:**
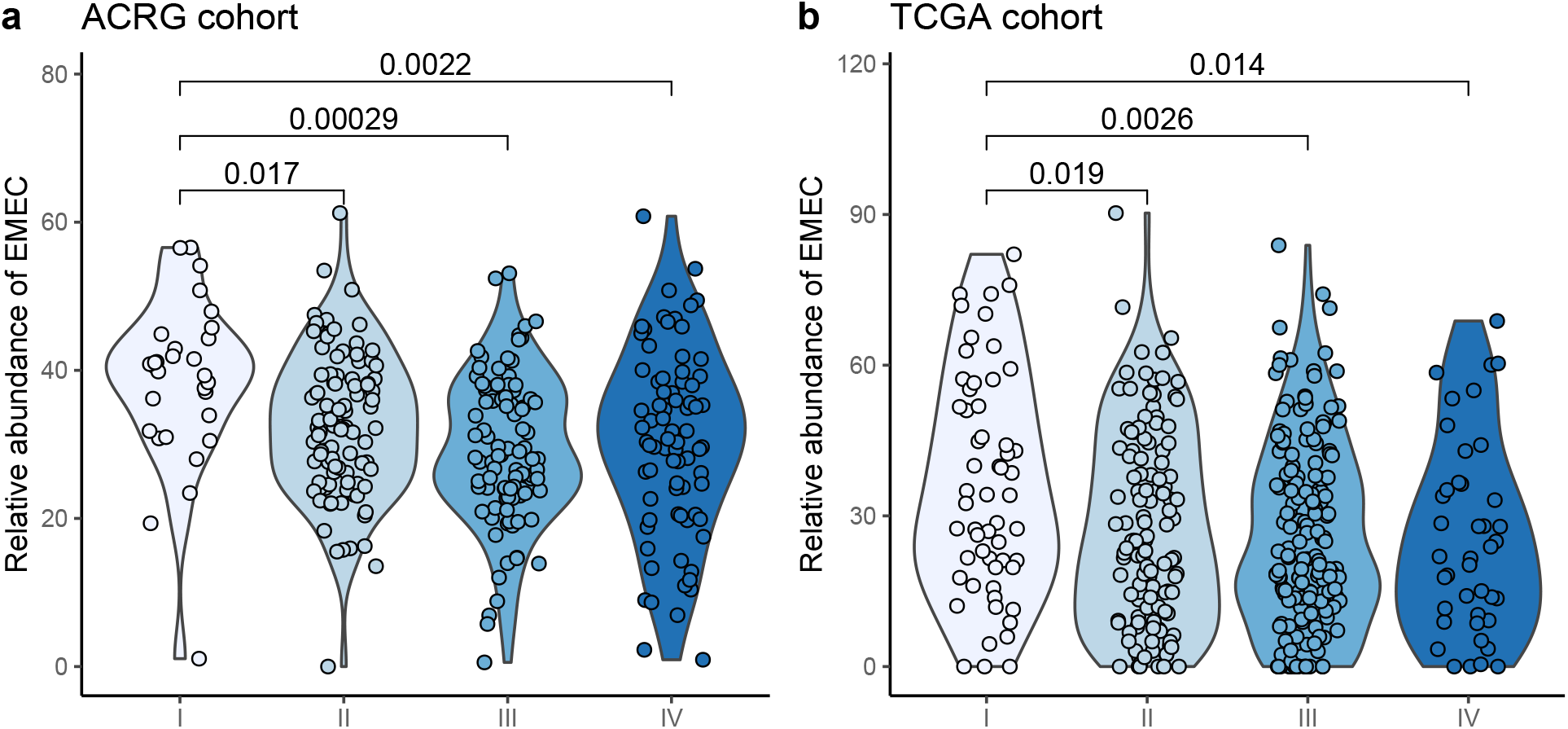
Comparison of the relative abundance of EMEC population across four stages in the ACRG and TCGA cohorts.

**Figure 6:**
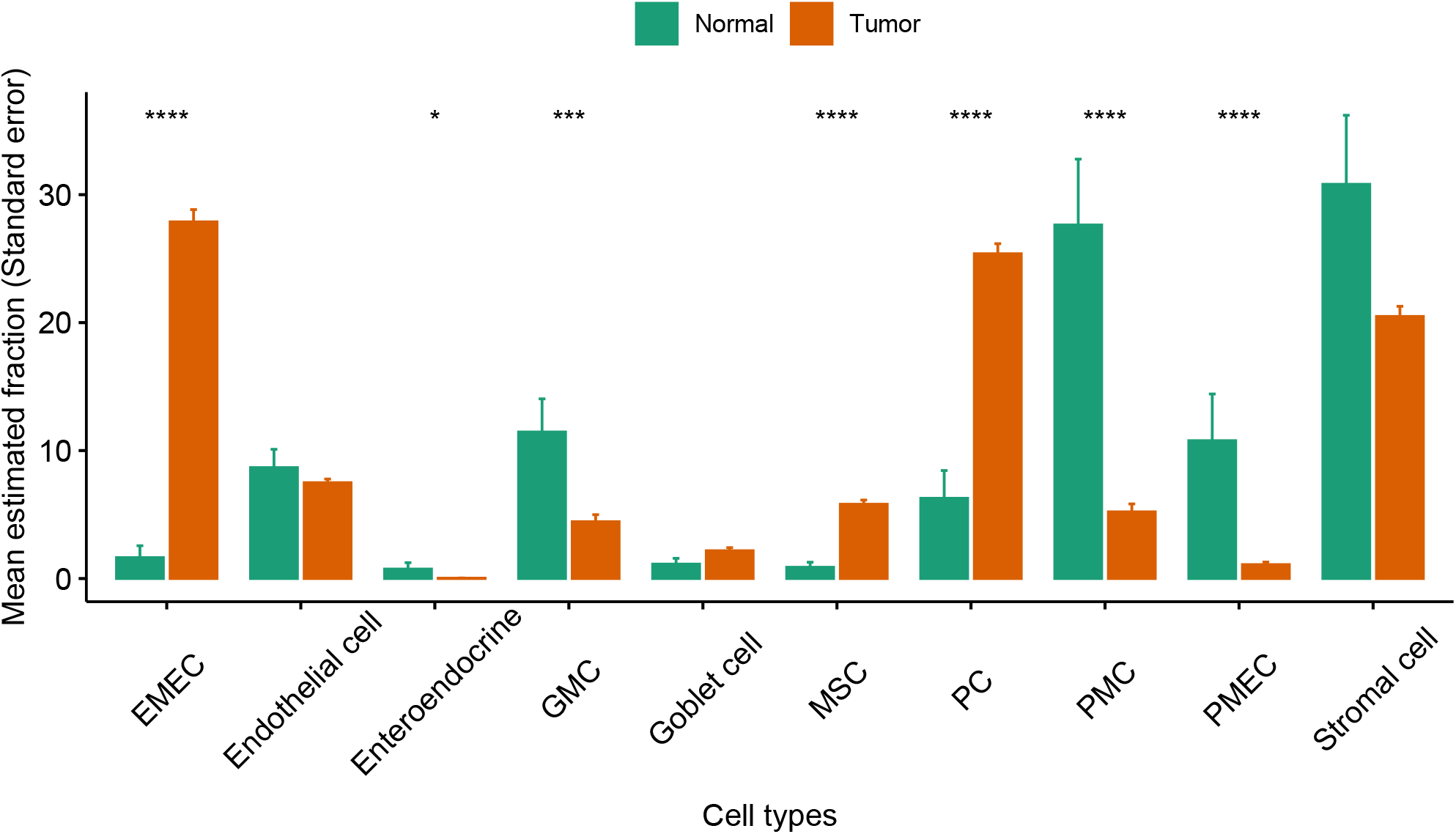
Comparison of the relative abundance of non-immune cell populations between normal and tumor samples in the TCGA-STAD cohort.

We also observed the PMEC population was significantly lower in tumor samples than in normal samples (*p* < 0.0001, Student’s t-test; Fig.6). The proliferative cell type was the second major epithelial cell type found in tumors. It was significantly increased in tumor samples when compared to normal samples (*p* < 0.0001, Student’s t-test; Fig.6). Contrastingly, the PMC cell types were significantly decreased in tumor samples compared to normal samples (*p* < 0.0001, Student’s t-test; Fig.6).

### 2.3 Correlates of non-immune/immune factors with overall survival

The ACRG (GSE62254) cohort was used as the training cohort because it provided the most comprehensive clinical data along with more than 5 years of follow-up information for 300 GC patients. The univariate Cox proportional hazards regression model was used to identify prognostic factors of overall survival (OS) in the training cohort. We further expanded the univariate analyses to Multivariate Cox proportional hazard analyses, which accounts for age, sex, stage, Lauren histology, and adjuvant chemotherapy treatment as additional clinical covariates to examine their independent prognostic values. The univariate Cox analyses indicated that the three non-immune factors of EMEC/stromal cell, PC/stromal cell, and endothelial cell/stromal cell ratios were significantly correlated to OS, and the hazard ratio ranged from 0.27 to 0.8 (*p* < 0.05). Additionally, the ratios of EMEC/stromal cell and PC/stromal cell remained significant prognostic factors of OS by multivariate analyses, with hazard ratios of 0.82 and 0.85 (*p* < 0.05), respectively (Table 1). The univariate Cox regression analysis for the prediction of OS also confirmed that three immune factors - the ratios of T adaptive/monocytes, monocytes/T adaptive, and monocytes/B adaptive were all found to be of prognostic significant factors with a hazard ratio ranging from 0.86 to 2.54 (*p* < 0.05). The multivariate analyses confirmed that the T adaptive/monocytes and monocytes/T adaptive ratios were independent prognostic factors of OS with hazard ratios of 0.88 and 2.61 (*p* < 0.05), respectively (Table 2).

**Table 1:**
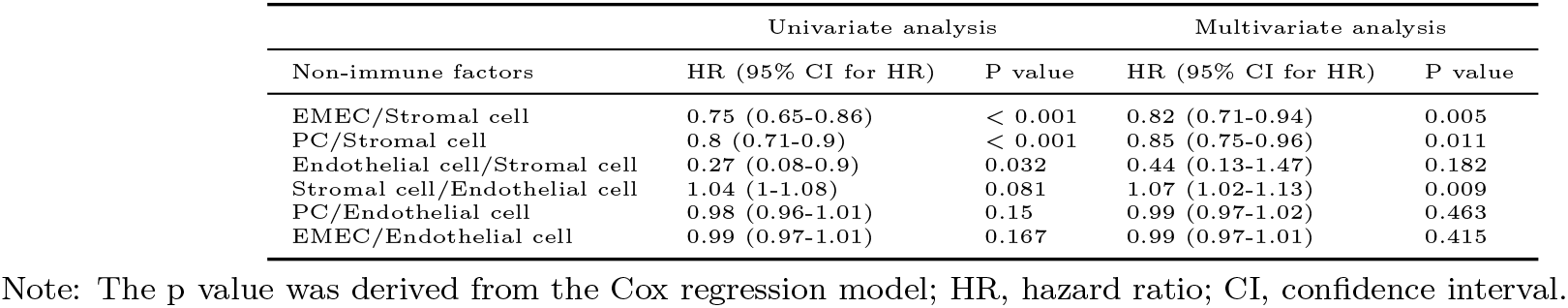
Univariate and multivariate results in Cox proportional hazards analysis of non-immune factors in the training cohort.

**Table 2:** Univariate and multivariate results in Cox proportional hazards analysis of immune factors in the training cohort.

| Immune factors | Univariate analysis |  | Multivariate analysis |  |
| --- | --- | --- | --- | --- |
|  | HR (95% CI for HR) | P value | HR (95% CI for HR) | P value |
| T adaptive/Monocytes | 0.86 (0.78-0.96) | 0.004 | 0.88 (0.8-0.97) | 0.01 |
| Monocytes/T adaptive | 2.54 (1.24-5.21) | 0.011 | 2.61 (1.11-6.15) | 0.028 |
| Monocytes/B adaptive | 1.14 (1-1.29) | 0.048 | 1.08 (0.95-1.22) | 0.239 |
| B adaptive/Monocytes | 0.8 (0.61-1.04) | 0.097 | 0.78 (0.59-1.03) | 0.078 |
| T Innate/T adaptive | 2.76 (0.81-9.39) | 0.104 | 1.6 (0.41-6.21) | 0.5 |
| Granulocytes/Monocytes | 0.79 (0.5-1.24) | 0.304 | 0.9 (0.63-1.29) | 0.582 |
| B adaptive/T Innate | 0.99 (0.96-1.02) | 0.347 | 0.99 (0.97-1.01) | 0.354 |
| T Innate/B adaptive | 1.16 (0.85-1.59) | 0.36 | 1 (0.72-1.37) | 0.978 |
| B adaptive/T adaptive | 1.44 (0.61-3.43) | 0.405 | 1.43 (0.54-3.78) | 0.471 |
| Granulocytes/B adaptive | 0.87 (0.63-1.22) | 0.426 | 1.05 (0.76-1.45) | 0.781 |
| Granulocytes/T Innate | 0.96 (0.86-1.09) | 0.553 | 1 (0.99-1.01) | 0.997 |
| T Innate/Monocytes | 0.89 (0.57-1.38) | 0.598 | 0.82 (0.52-1.3) | 0.393 |
| T adaptive/B adaptive | 0.99 (0.93-1.05) | 0.742 | 0.99 (0.93-1.06) | 0.817 |
| Granulocytes/T adaptive | 0.85 (0.32-2.29) | 0.749 | 1.17 (0.46-2.98) | 0.741 |

Thus, we defined a new TME score, called STEM score for each GC sample by combining the most significant non-immune (the ratio of EMECs to Stromal cells) and immune factors (the ratio of adaptive T cells to Monocytes) as:

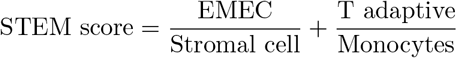

We performed multivariate Cox regression analysis correcting for clinicopathological variables, including age, sex, stage, Lauren histology, and adjuvant chemotherapy treatment. We found that the STEM score acts as an independent prognostic factor for OS (HR 0.9, *p* = 0.003) in the training cohort (Table 3).

**Table 3:**
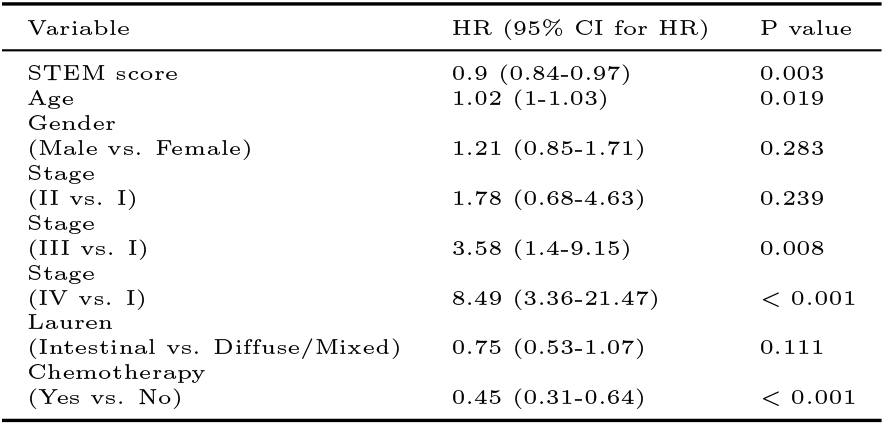
Multivariate Cox regression analysis used to evaluate the independent risk factor of prognosis in the training cohort.

| Variable | HR (95% CI for HR) | P value |
| --- | --- | --- |
| STEM score | 0.9 (0.84-0.97) | 0.003 |
| Age | 1.02 (1-1.03) | 0.019 |
| Gender<br>(Male vs. Female) | 1.21 (0.85-1.71) | 0.283 |
| Stage<br>(II vs. I) | 1.78 (0.68-4.63) | 0.239 |
| Stage<br>(III vs. I) | 3.58 (1.4-9.15) | 0.008 |
| Stage<br>(IV vs. I) | 8.49 (3.36-21.47) | < 0.001 |
| Lauren<br>(Intestinal vs. Diffuse/Mixed) | 0.75 (0.53-1.07) | 0.111 |
| Chemotherapy<br>(Yes vs. No) | 0.45 (0.31-0.64) | < 0.001 |

To validate whether the STEM score had consistent prognostic value in different cohorts, we applied it to three independent validation data sets from TCGA-STAD, GSE15459, and GSE84437. The multivariate Cox proportional hazard analyses, which account for age, sex, stage, Lauren histology, and adjuvant chemotherapy/radiation therapy treatment (if applicable as additional clinical covariates) confirmed that the STEM score was an independent prognostic factor of OS in each validation data set (TCGA-STAD: HR 0.94, *p* = 0.001, GSE15459: HR 0.89, *p* = 0.01, and GSE84437: HR 0.86, *p* < 0.001). We further performed a meta-analysis to evaluate the overall effect of the STEM score on clinicopathologic factor-adjusted survival. We added three validation cohorts to the training cohort. A forest plot of estimated hazard ratios indicated the STEM score was a significant independent prognostic factor of OS (HR = 0.92, *p* = 2.05 × 10^−9^, z-test for overall effect; Fig.7).

**Figure 7:**
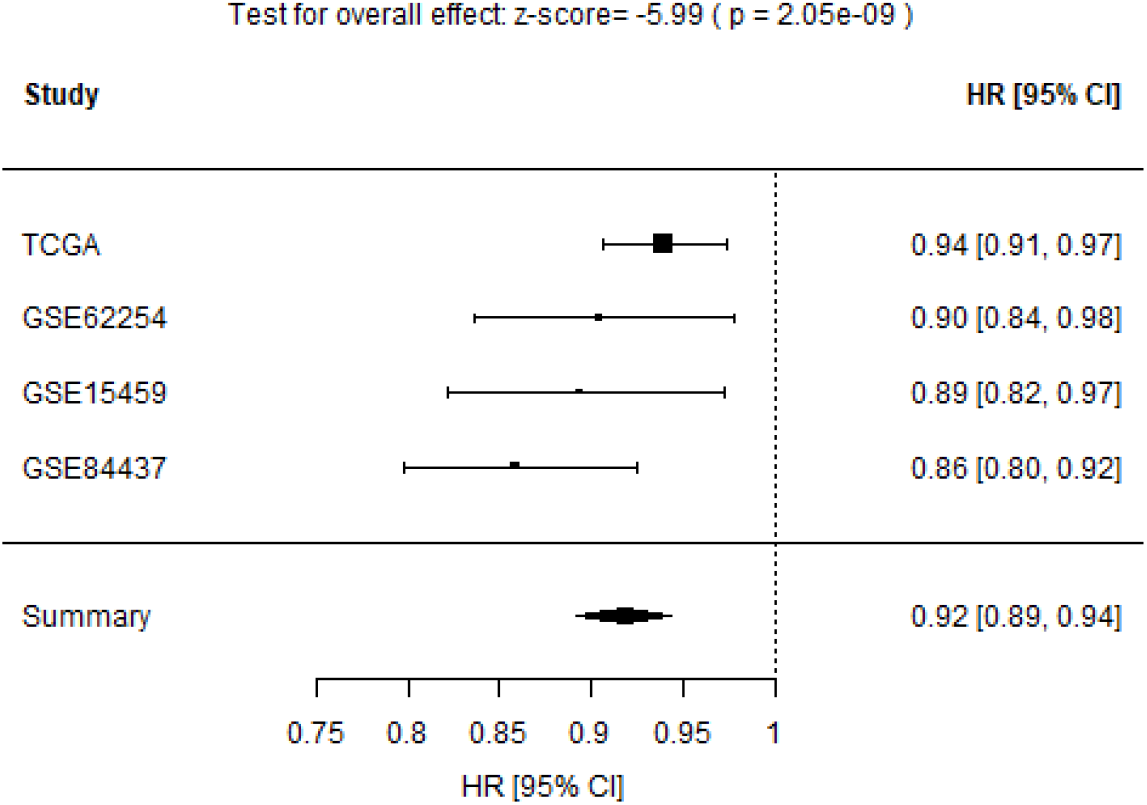
Forest plot of estimated hazard ratios indicating STEM score as a prognostic factor for OS in four GC cohorts.

### 2.4 Increased STEM score associated with superior survival

We next assessed the predictive value of the STEM score for risk stratification. For the ACRG cohort, we stratified 300 patients into two groups (low vs. high) according to the STEM score and using an optimal cutoff of 3.95 (Fig.8; see Methods for details). The Kaplan-Meier curve showed that patients in the high-STEM score group had significantly longer OS times than patients in the low-STEM score group (*p* < 0.0001, log-rank test; Fig.9).

**Figure 8:**
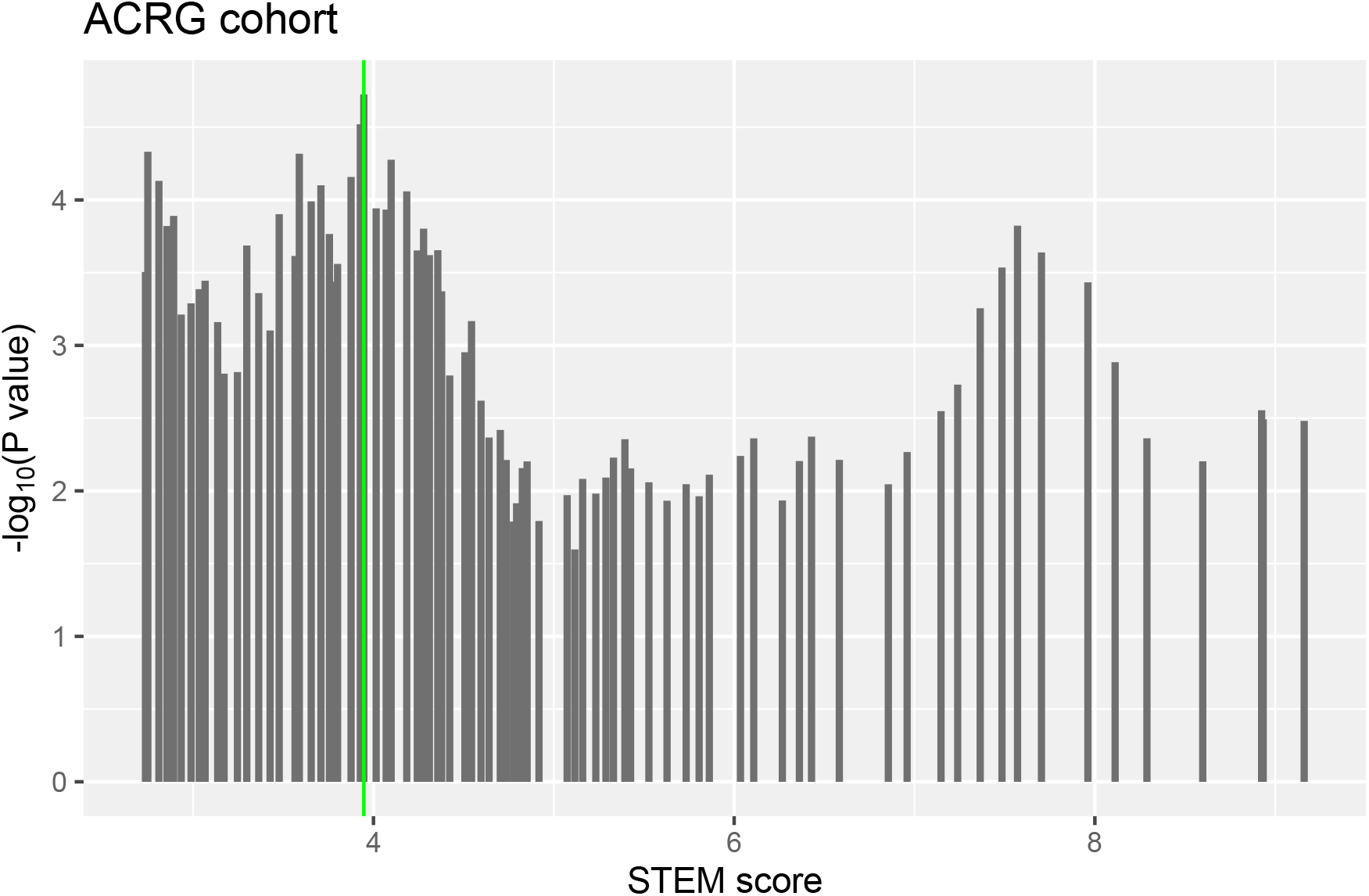
An illustration of optimal cutoff identification for STEM score.

**Figure 9:**
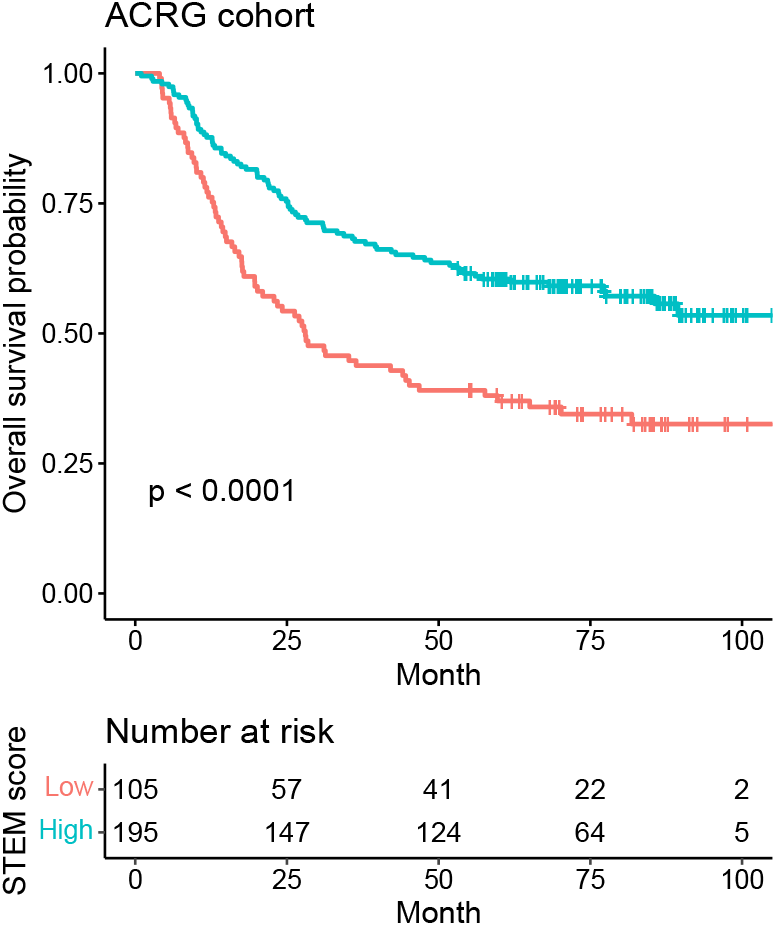
Kaplan-Meier survival curves for the patients with low vs. high STEM scores in the ACRG cohort.

By using the same cutoff optimized in the ACRG cohort, we observed consistent results across three independent validation cohorts that showed patients with a high STEM score yielded better OS than those with a low STEM score (GSE15459: *p* = 0.015; GSE84437: *p* = 4 × 10^−4^; TCGA-STAD: *p* = 1.8 × 10^−4^; a combined set of four cohorts: *p* < 0.0001; log-rank test, Fig.10).

**Figure 10:**
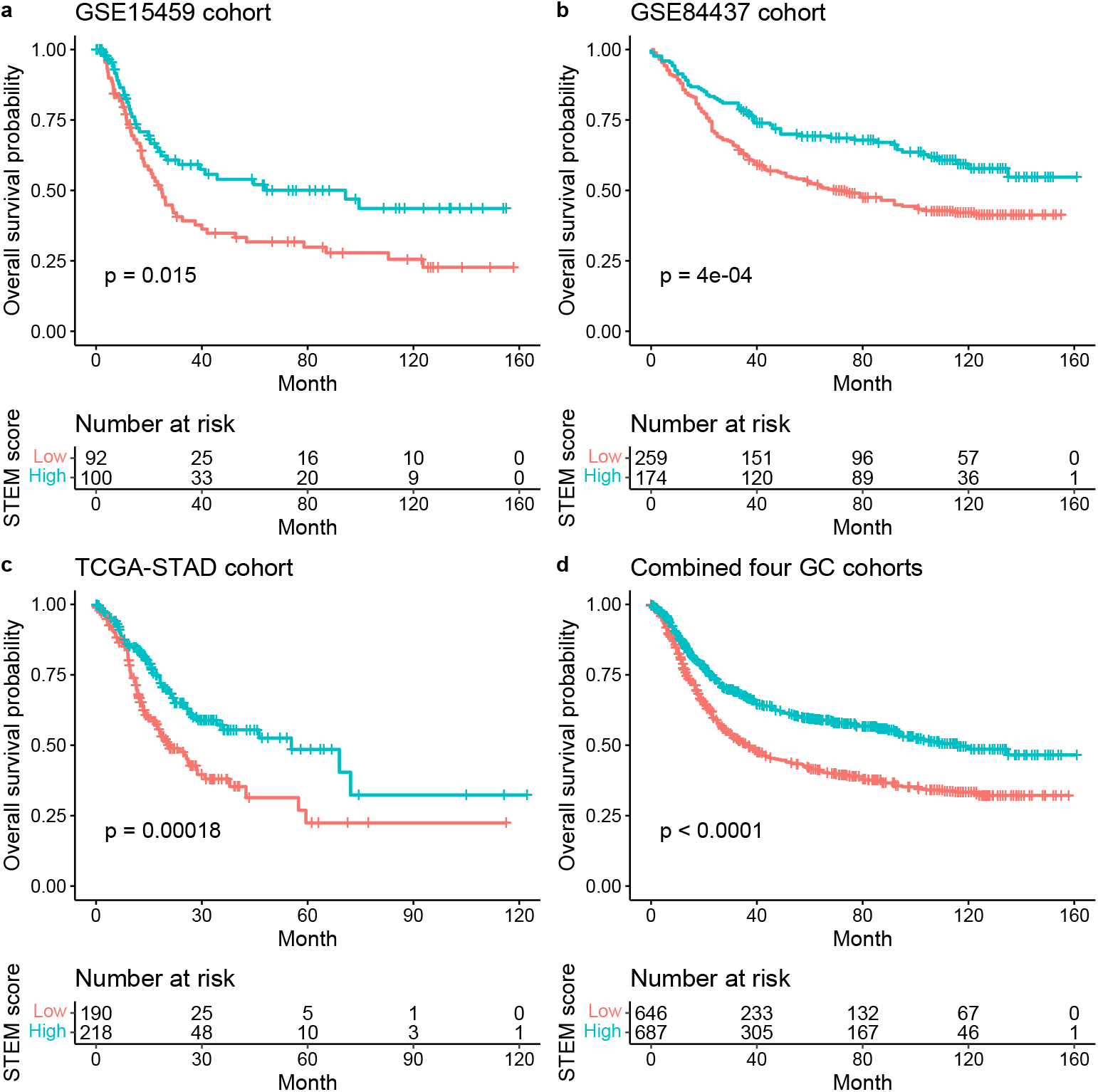
Kaplan-Meier plots for the OS of patients with low vs. high STEM scores in the GSE15459 (a), GSE84437 (b), TCGA-STAD (c) cohorts and a combined set of four cohorts (ACRG, GSE15459, GSE84437, and TCGA-STAD) (d). Significance test p value is shown in the lower left.

Additionally, we investigated the prognostic values of the STEM score in groups of patients treated with or without chemotherapy. We then stratified the ACRG cohort into four groups based on the STEM score and chemotherapy (CT) treatment or not. The unadjusted survival curve for the four groups indicated the high-STEM score groups had superior survival compared to the low-STEM score group for patients regardless of chemotherapy treatment (Fig.11a).

**Figure 11:**
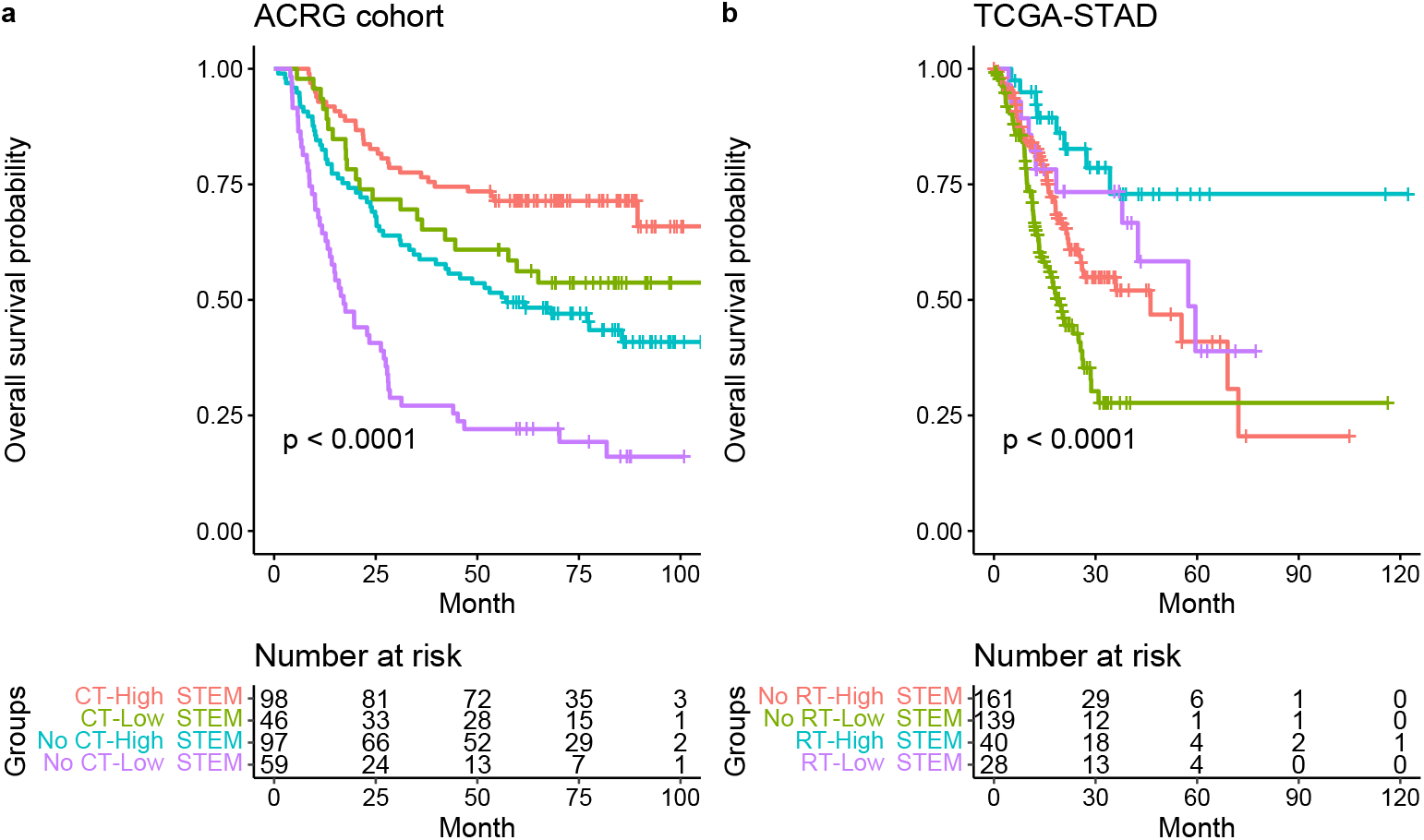
(a) Kaplan-Meier survival curves for the four groups (including chemotherapy-high STEM score, chemotherapy-low STEM score, no chemotherapy-high STEM score, and no chemotherapy-low STEM score) in the ACRG cohort. Significance test p value is shown in the lower left. (b) Kaplan-Meier survival curves for the four groups (including radiation therapy-high STEM score, radiation therapy-low STEM score, no radiation therapy-high STEM score, and no radiation therapy-low STEM score) in the TCGA-STAD cohort.

To explore the prognostic values of the STEM score in groups of patients treated with or without radiation therapy, we stratified the TCGA-STAD cohort into four groups based on the STEM score and radiation therapy (RT) treatment or not. The unadjusted survival curve for the four groups indicated the high-STEM score groups still had superior survival compared to the low-STEM score group for patients regardless of radiation therapy treatment (Fig.11b).

By using multivariate Cox regression analysis and univariate Kaplan-Meier analysis in Section 2.3, four prognostic cell populations of Stromal cell, adaptive T cell, EMEC and Monocyte, referred as STEM populations, that had been found to be significant in the univariate Cox regression analyses were further examined to be prognosis stratification factors in gastric cancer. Thus, these four cell populations were selected to perform cluster analysis of all 1340 GC samples in four GC cohorts. Based on the estimated relative abundance of these four cell populations, we applied spectral clustering with the optimal number of clusters chosen by NbClust (see Methods for details). The three resulting TME subtypes, including TMEsubtype-H, TMEsubtype-M, and TMEsubtype-L (with 302, 591, and 447 samples, respectively) were characterized by a distinct distribution of relative abundance over the four selected cell populations. The relative abundance of these four cell populations varied significantly across the three TME subtypes (*p* < 2.22 × 10^−16^, Kruskal-Wallis test; Fig.12). The TMEsubtype-H cluster had the highest stromal cell (mean proportion = 0.48, *p* < 2.22 × 10^−16^, Wilcoxon test relative to next-highest; Fig.12, left bottom) and monocyte abundance (mean proportion = 0.29, *p* < 2.22 × 10^−16^, Wilcoxon test relative to next-highest; Fig.12, right bottom), while it had the lowest EMEC (mean proportion = 0.17, *p* < 2.22 × 10^−16^, Wilcoxon test relative to next-lowest; Fig.12, left top) and adaptive T cell abundance (mean proportion = 0.44, *p* = 0.0036, Wilcoxon test relative to next-lowest; Fig.12, right top). However, in the TMEsubtype-L the opposite was observed. The TMEsubtype-L cluster was found to have the lowest stromal cell (mean proportion = 0.14, *p* < 2.22 × 10^−16^, Wilcoxon test relative to next-lowest; Fig.12, left bottom) and monocyte abundance (mean proportion = 0.18, *p* = 6.7 × 10^−9^, Wilcoxon test relative to next-lowest; Fig.12, right bottom), while it had the highest EMEC (mean proportion = 0.44, *p* < 2.22 × 10^−16^, Wilcoxon test relative to next-highest; Fig.12, left top) and adaptive T cell abundance (mean proportion = 0.57, *p* = 0.0036, Wilcoxon test relative to next-highest; Fig.12, right top). The mean proportions of EMEC, stromal cell, adaptive T cell, and monocyte for the TMEsubtype-M were 0.25, 0.22, 0.47, and 0.2, respectively.

**Figure 12:**
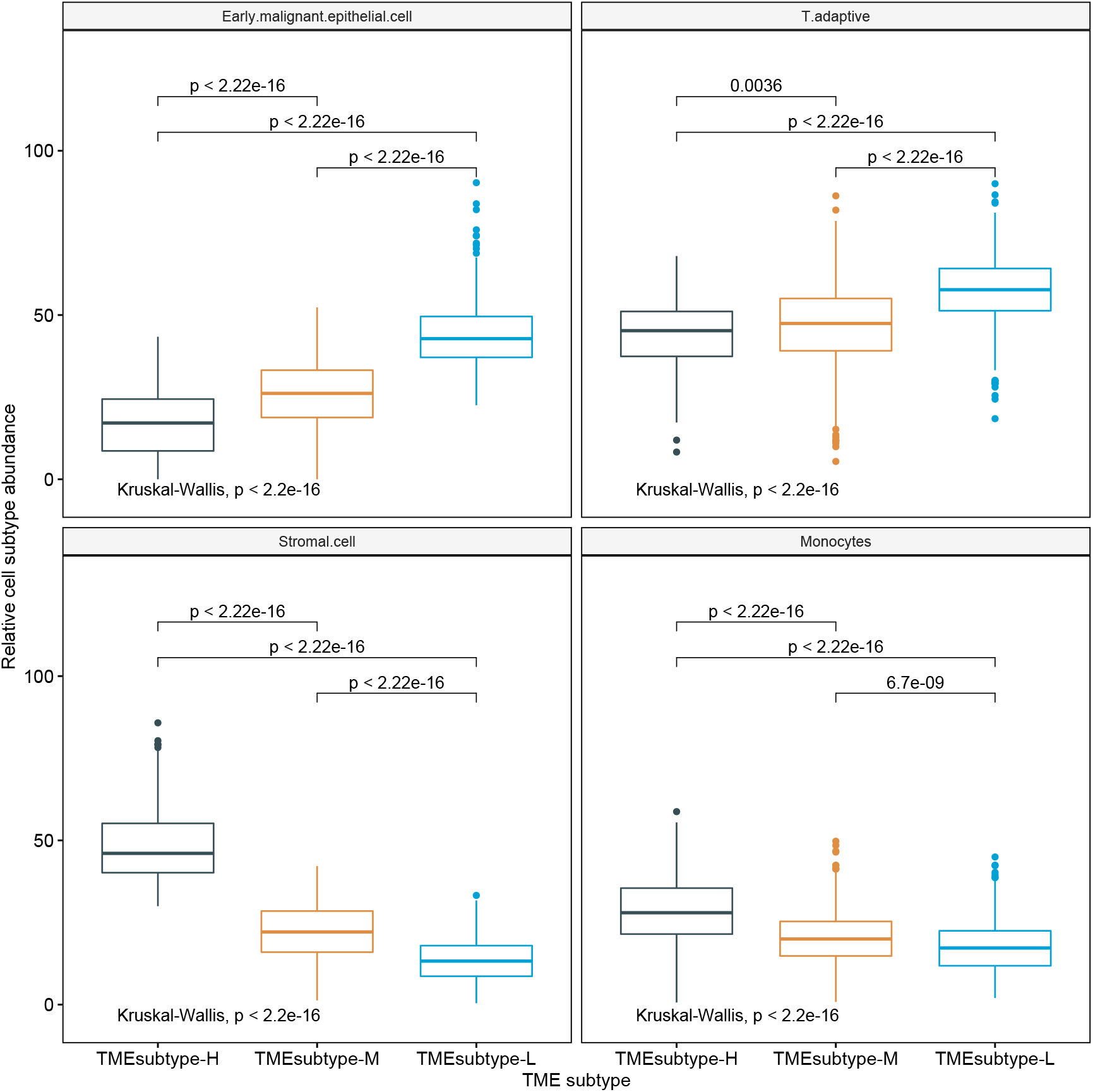
Comparison of the estimated proportions of four prognostic cell populations across three TME subtypes in the meta cohort.

We further examined the TME subtypes’ association with OS. The TMEsubtype-L had the best prognosis (OS HR (95% CI) 0.52 (0.42-0.65), *p* < 0.0001 relative to the TMEsubtype-H, adjusted for age and sex), the TMEsubtype-M had intermediate prognosis (OS HR (95% CI) 0.76 (0.63-0.91), *p* = 0.0035 relative to the TMEsubtype-H, adjusted for age and sex), and the TMEsubtype-H had the least favorable outcome (*p* < 0.0001, log-rank test; Fig.13a). A decreased value of the STEM score, non-immune, or immune score led to worse outcomes in the TMEsubtype-H (Fig.13b). The survival analysis revealed a substantial difference in OS among the three TME subtypes. Robust correlations between the identified TME subtypes and OS were also validated in ACRG (*p* < 0.0001, log-rank test; Fig.14a), GSE84437 (*p* = 0.002, log-rank test; Fig.14b), and TCGA-STAD (*p* = 0.014, log-rank test; Fig.14c) cohorts, separately. Patients in different GC cohort were stratified into three groups with significantly distinct prognosis, and it was found that the lower the STEM score the poorer the patient survival outcome. Although the difference in the GSE15459 cohort was not statistically significant (*p* = 0.097, log-rank test; Fig.14d), the TMEsubtyp-H still had the poorest outcome.

**Figure 13:**
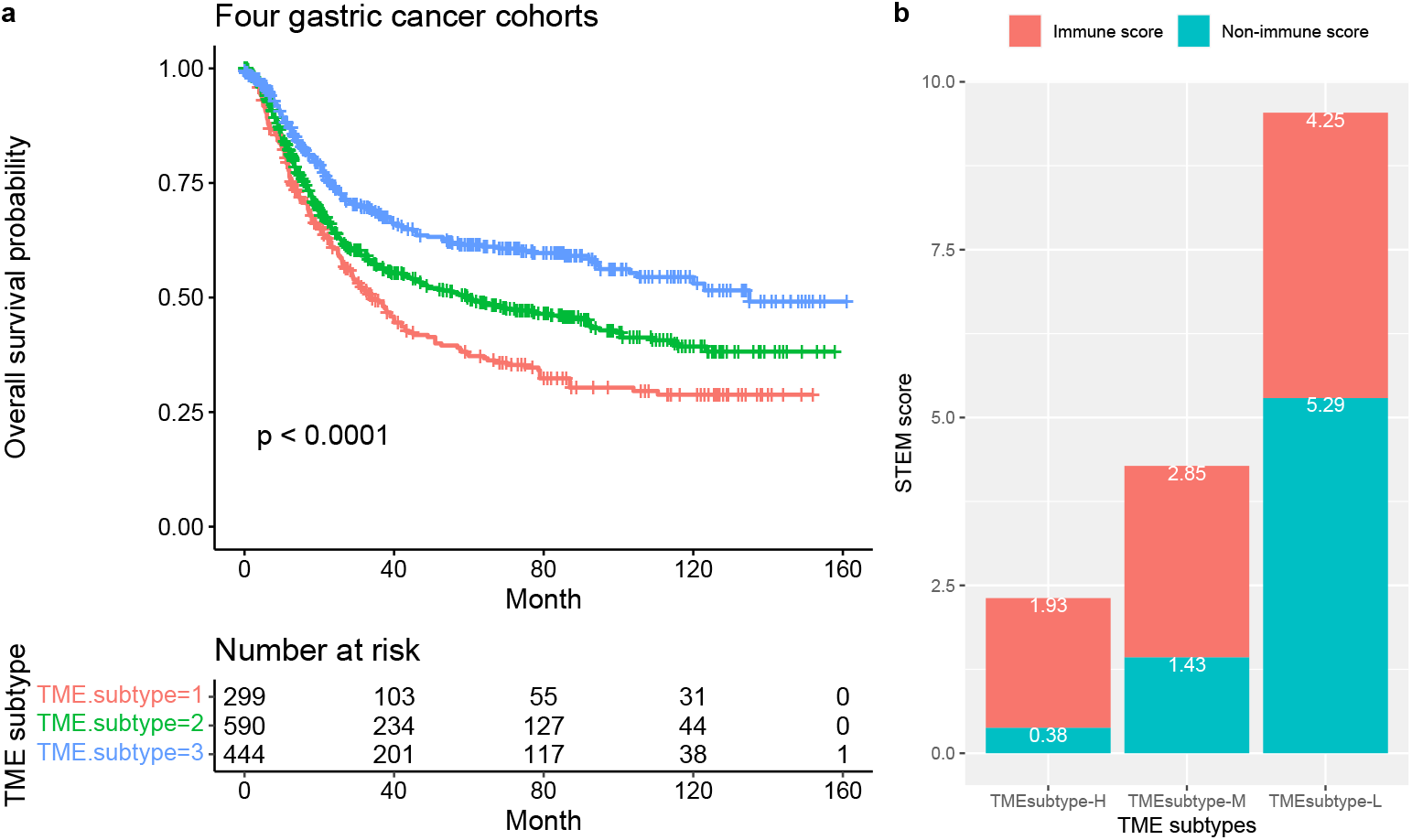
TME subtype in gastric cancer. (a)Kaplan-Meier plots for the OS of patients stratified into three TME subtypes in the meta cohort (ACRG, GSE15459, GSE84437 and TCGA-STAD). Significance test p value is shown in the lower left. (b) The mean non-immune and immune scores with TME subtypes.

**Figure 14:**
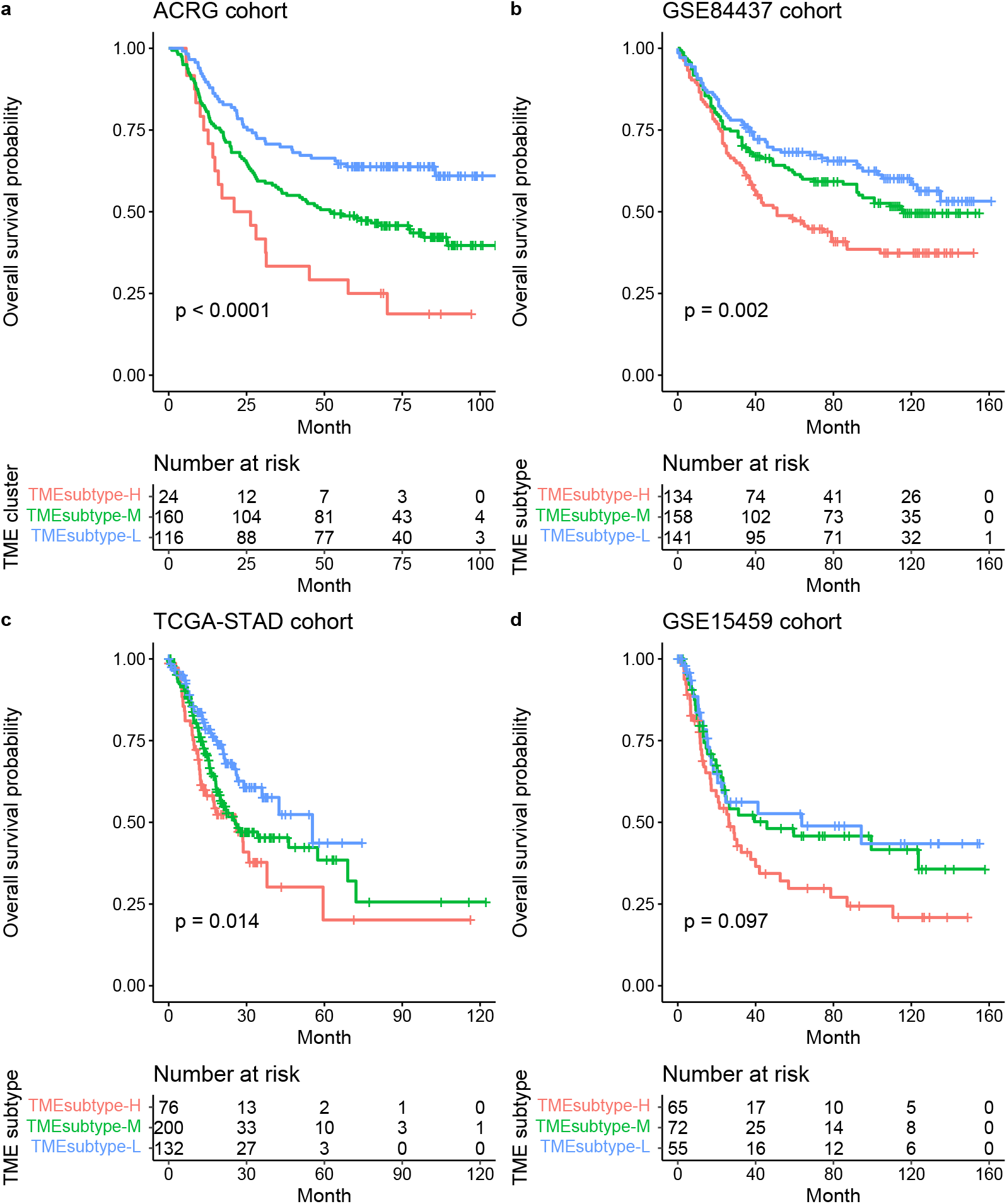
Kaplan-Meier plots for the OS of patients stratified into three TME subtypes in the ACRG (a), GSE84437 (b), TCGA-STAD (c) and GSE15459 (d) cohorts. Significance test p value is shown in the lower left.

The risk stratification model was further investigated in patients with the same stage GC in the ACRG and TCGA-STAD data sets. Similarly to the whole cohort, patients with stage IV GC were stratified into three groups with significantly distinct prognosis (ACRG: *p* = 0.031, Fig.15a and TCGA-STAD: *p* = 0.028, Fig.15b; log-rank test).

**Figure 15:**
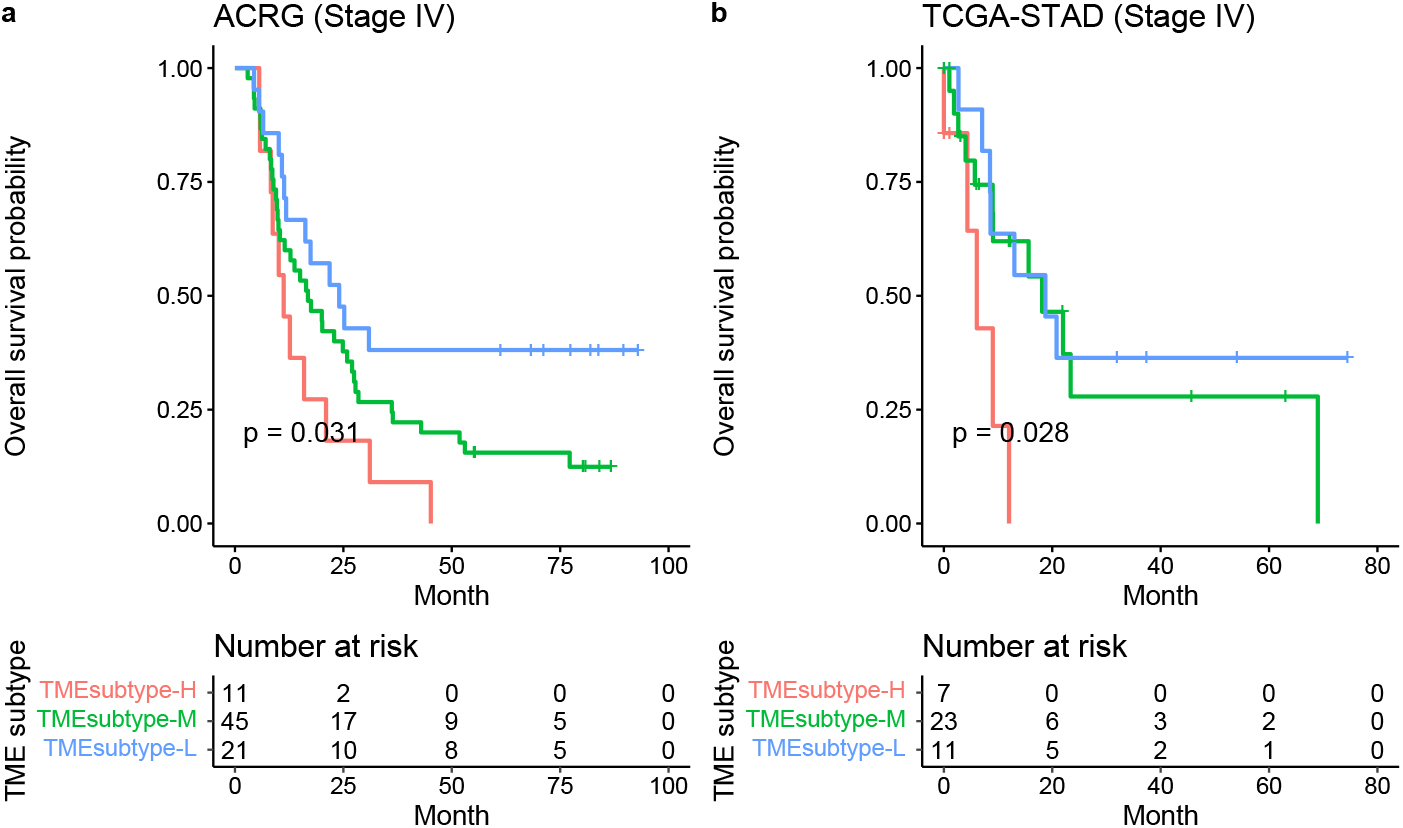
TME subtype in patients with stage IV GC. Kaplan-Meier plots for the OS of patients with stage IV GC stratified into three TME subtypes in the ACRG (a) and TCGA-STAD (b) cohorts.

To investigate the predictive value of the TME subtypes for radiation therapy response in the TCGA-STAD cohort, we evaluated the association between TME subtypes and overal survival among stage III patients who either received or did not receive radiation therapy. We found that for patients in the TMEsubtype-M and TMEsubtype-L group, radiation therapy was associated with improved OS (HR 0.16, 95% CI (0.06-0.4), *p* < 0.0001; Fig.16a). However, for patients in the TMEsubtype-H group, performing radiation therapy did not improve OS (HR 0.49, 95% CI (0.14-1.72), *p* = 0.25; Fig.16b). The mean STEM score of patients in the TMEsubtype-H group is substantially smaller than those in the TMEsubtype-M and TMEsubtype-L groups (Fig.16c).

**Figure 16:**
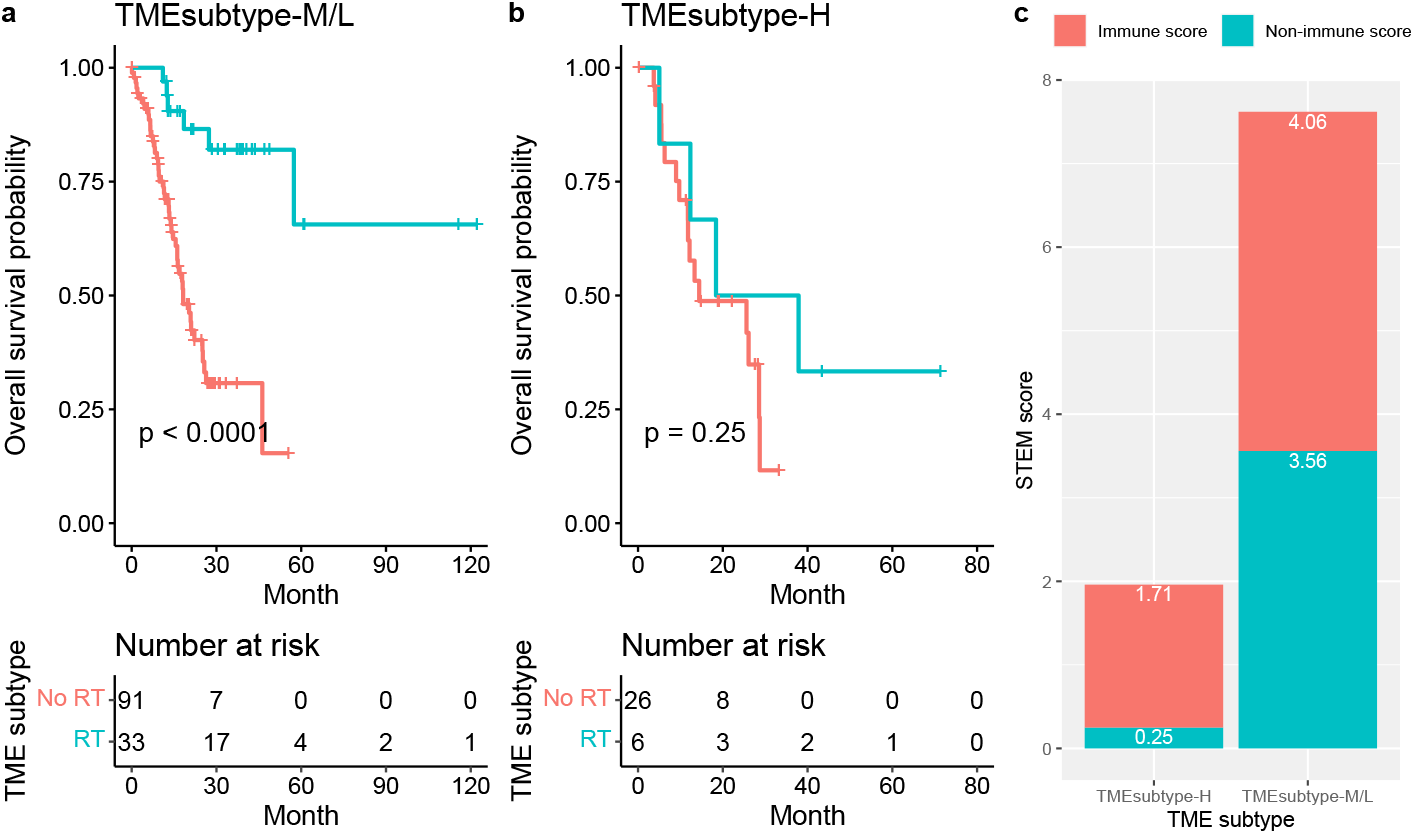
Relationship between the TME subtypes and survival benefit from radiation therapy in matched patients with stage III gastric cancer: (a) TMEsubtype-H, (b) TMEsubtype-M/L. (c) Compare the mean non-immune and immune scores of patients in the TMEsubtype-H to the TMEsubtype-M and -L for stage III disease in the TCGA-STAD cohort.

### 2.5 Comparison with other reported molecular classifications for GC

We compared the similarities and differences of the identified TME subtypes with the molecular subtypes derived by the ACRG, as well as with the genomic subtypes derived by the TCGA. The ACRG classified GC into four molecular subtypes, including EMT, MSS/TP53-, MSS/TP53+, and MSI, which are associated with distinct molecular alterations, disease progression, and survival outcomes based on gene expression data. In survival analysis, the MSI subtype had the best prognosis, followed by MSS/TP53+, MSS/TP53-, and finally the EMT subtype having the worst prognosis [4]. The comparisons of the TME subtypes with the ACRG molecular subtypes showed several differences, for instance, the samples classified as the ACRG EMT subtype were present in both the TMEsubtype-H and TMEsubtype-M, and the samples classified as ACRG MSS (TP53+ and TP53-) and MSI subtypes were present in both the TMEsubtype-M and TMEsubtype-L. However, we observed that the TMEsubtype-H, TMEsubtype-M, and TMEsubtype-L were enriched in ACRG EMT, MSS (TP53+ and TP53-) and MSI, respectively (Fig.17a). Moreover, we investigated the association of the TME subtypes with tumor stages. The TMEsubtype-H was linked to patients classified as stage III and IV, whereas the TMEsubype-L was associated with patients classified as early stage I and II (Fig.17a). We showed previously that a lower STEM score was associated with poorer survival outcome. We found similar results when comparing the STEM score across ACRG molecular subtypes. The ACRG

EMT subtype, composed mostly of diffuse-type tumors, had been shown to have the worse prognosis of the four and was linked to the lowest STEM score (mean STEM score = 3.1, *p* = 2.3 × 10^−14^, Wilcoxon test relative to next-lowest). The ACRG MSI subtype, which had been shown to have the best prognosis of four, was linked to the highest STEM score (mean STEM score = 6.49, *p* = 0.0073, Wilcoxon test relative to next-highest). The mean STEM score of the ACRG MSS subtype was 5.73. The STEM score varied significantly across the four ACRG molecular subtypes (*p* = 3.1 × 10^−16^, Kruskal-Wallis test; Figure.17b). Consistent results were found for the non-immune and immune scores across the ACRG molecular subtypes (Figure.17c).

**Figure 17:**
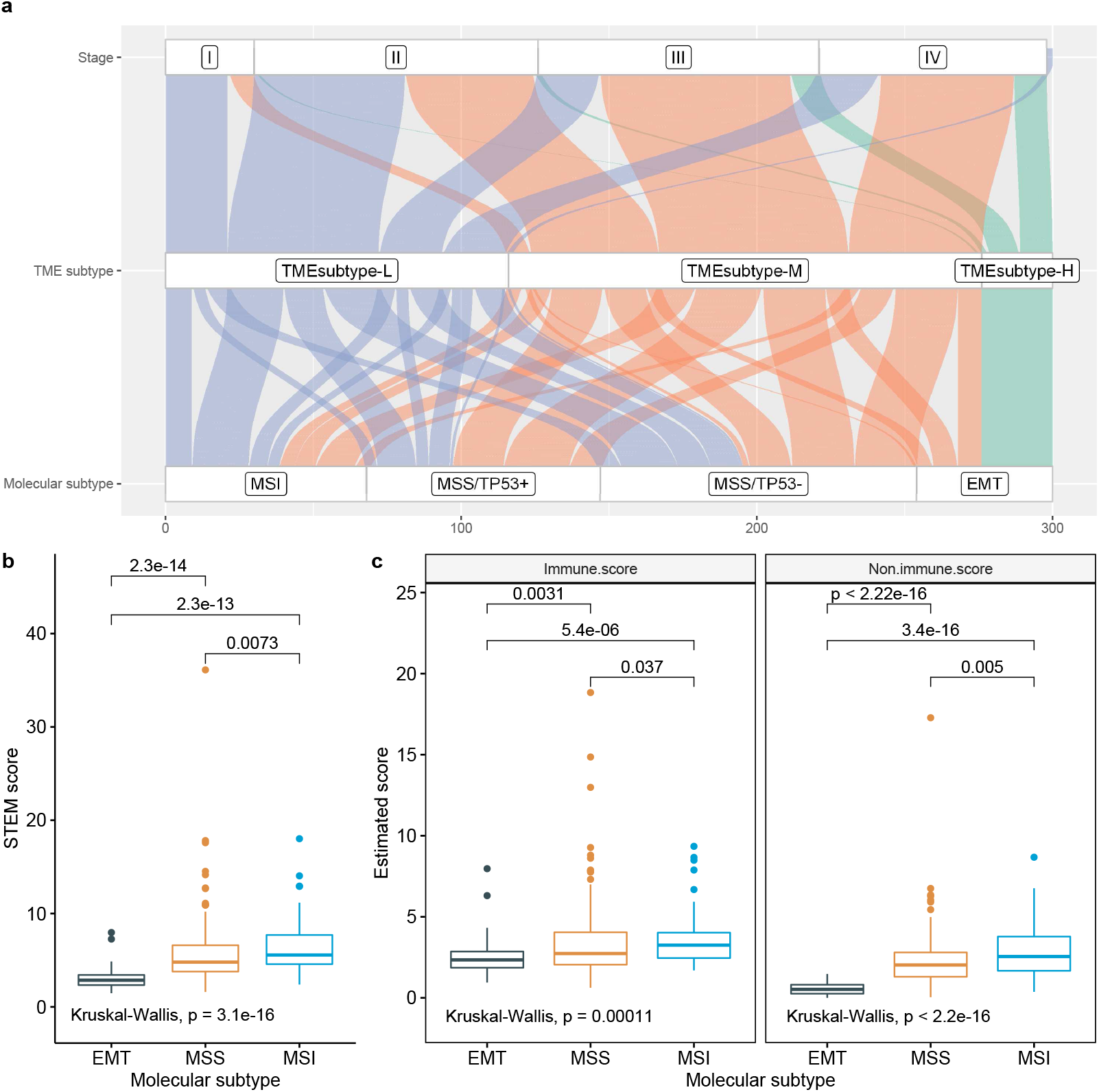
Association of TME subtypes and ACRG molecular subtypes. (a) Alluvial plot displaying the association of TME subtypes, ACRG molecular subtype and stage. (b) Comparison of STEM score across three ACRG molecular subtypes. (c) Comparison of non-immune (e.g. ratio of EMEC/stromal cell) and immune scores (e.g. ratio of T adaptive/monocytes) across three ACRG molecular subtypes.

The TCGA research network classified GC into four genomic subtypes, including EBV, MSI, GS, and CIN, by integrating data from six molecular platforms and performing Microsatellite instability (MSI) testing [5]. The MSI and EBV subtypes were shown to have a better prognosis than GS and CIN subtypes [4, 29, 30]. The comparisons of the TME subtypes with the TCGA genomic subtypes showed similarities, such as the TMEsubtype-H, TMEsubtype-M, and TMEsubtype-L were enriched in TCGA GS, CIN and MSI, respectively (Fig.18a). The TMEsubtype-H is primarily composed of samples classified as the TCGA GS and CIN subtypes. The GC samples classified as TMEsubtype-M were present across all TCGA genomic subtypes. The TMEsubtype-L is primarily composed of samples classified as the TCGA MSI, EBV, and CIN subtypes. We found a significantly lower mean STEM score in the TCGA GS (3.57) and CIN (5.7) subtypes compared to the EBV (9.41) and MSI (11.4) subtypes (Fig.18b), further reinforcing the prognostic value of the STEM score.

**Figure 18:**
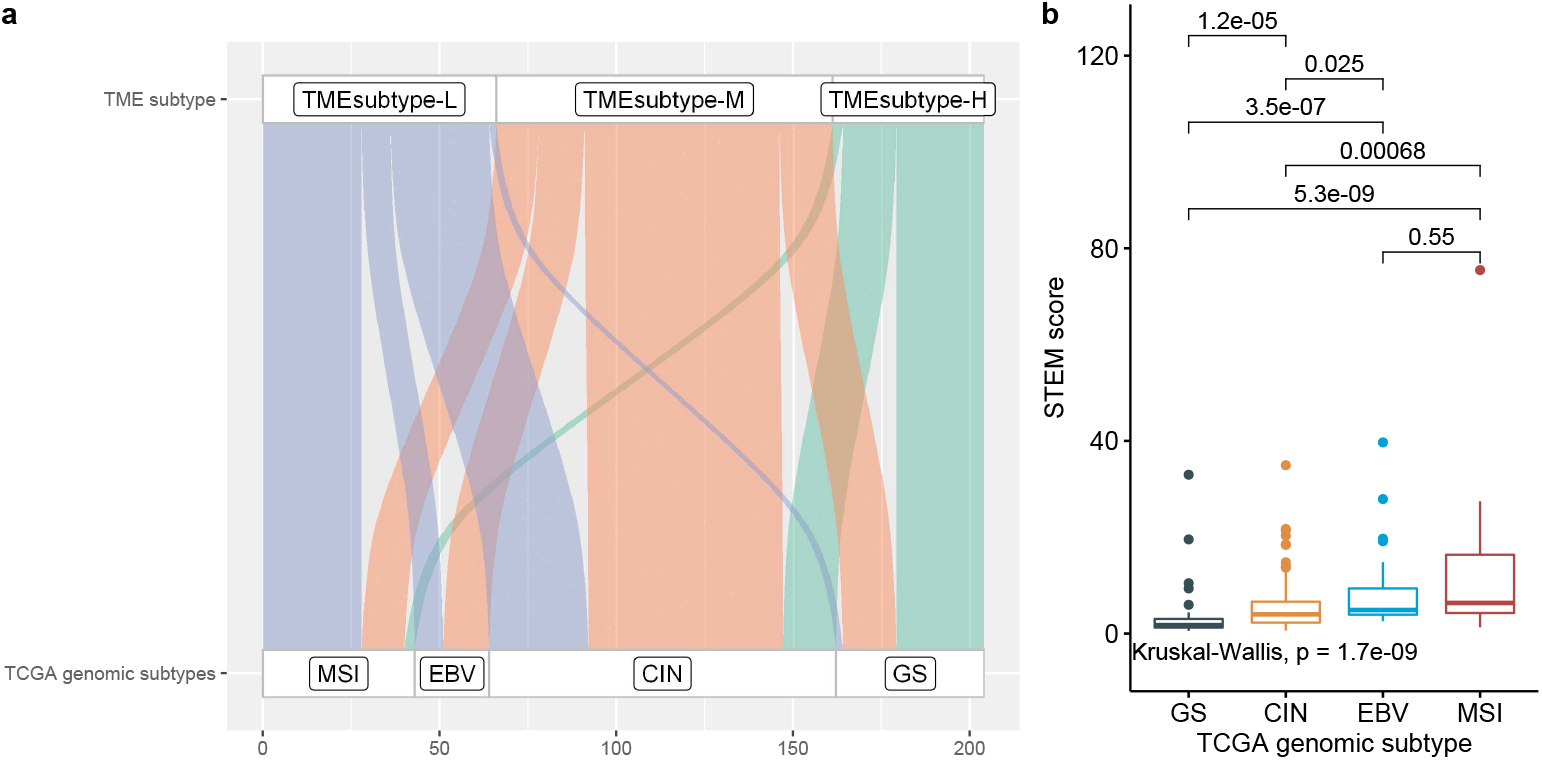
Association of TME subtypes and TCGA genomic subtypes. (a) Alluvial plot displaying the association of TME subtypes and TCGA-STAD subtypes. (b) Comparison of STEM score across four TCGA genomic subtypes.

### 2.6 Identification of GC prognostic gene signatures

We have shown predictive values of the EMEC and stromal cell populations for OS. In this respect, we further investigated prognostic gene signatures of these two cell populations. By using scRNA-seq transcription profiles, we have identified 150 gene markers for the EMEC and stromal cell populations on the construction of non-immune signature matrix. For each marker gene, we did multivariate Cox proportional hazard modeling on the four GC cohorts, respectively, accounting for conventional clinical and pathologic factors including age, sex, stage, Lauren histology, and adjuvant chemotherapy/radiation therapy treatment if applicable. Next, we performed fixed effects meta-analyses to identify gene signatures whose expressions were significantly associated with survival outcome across multiple cohorts.

A higher abundance of the stromal cell subtype has been associated with poorer prognosis. Thus, we identified those prognostic genes for each GC cohort at a p-value cutoff of 0.05, as well as those with a hazard ratio greater than 1, which suggest a significant increase in risk. There were 43 significant prognostic gene signatures detected after performing meta-analysis (*p* < 10^−5^, z-test for overall effect). The top 8 genes, including FERMT2 (Fermitin family homolog 2, HR=1.49), SGCE (sarcoglycan epsilon, HR=1.5), PPP1R14A (protein phosphatase 1 regulatory inhibitor subunit 14A, HR=1.38), LAMC1 (laminin subunit gamma 1, HR=1.83), MYL9 (myosin light chain 9, HR=1.3), TPM2 (tropomyosin 2, HR=1.34), TAGLN (transgelin, HR=1.33), and AKAP12 (A-kinase anchoring protein 12, HR=1.4), that have a smaller p value for overall effect are shown in Fig.19. We further explored the association between the expression level in prognostic marker genes with TME subtypes. We focused on the ACRG cohort. Violin plots indicate that the representative prognostic marker genes were highly expressed in TMEsubtype-H with relatively low levels in other two TME subtypes: TMEsubtype-M and TMEsubtype-L (Fig.20). The expression levels were significantly different among three TME subtypes (*p* < 10^−21^, one-way ANOVA F test).

**Figure 19:**
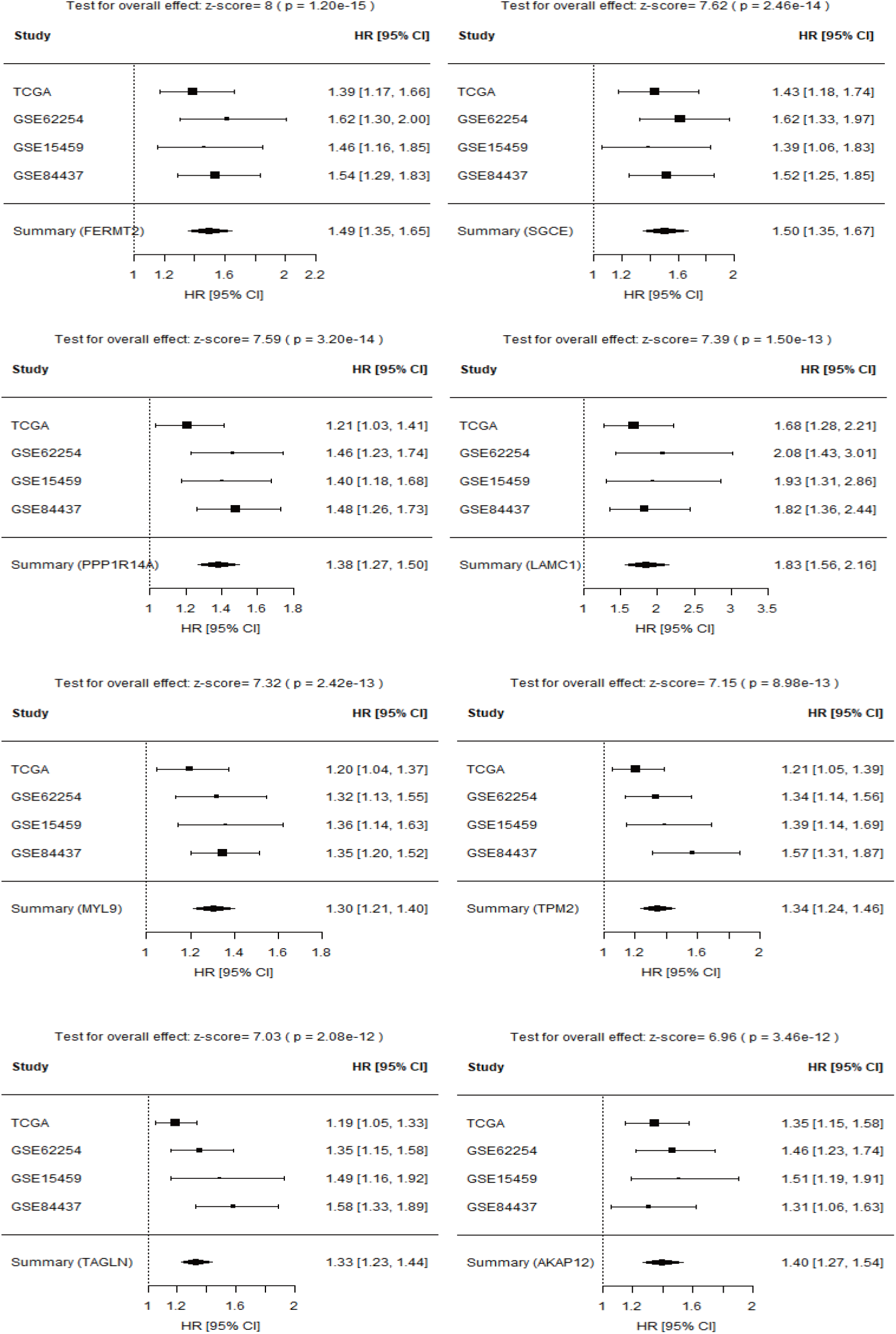
Forest plot of estimated hazard ratios of prognostic gene signatures in stromal cell subtype.

**Figure 20:**
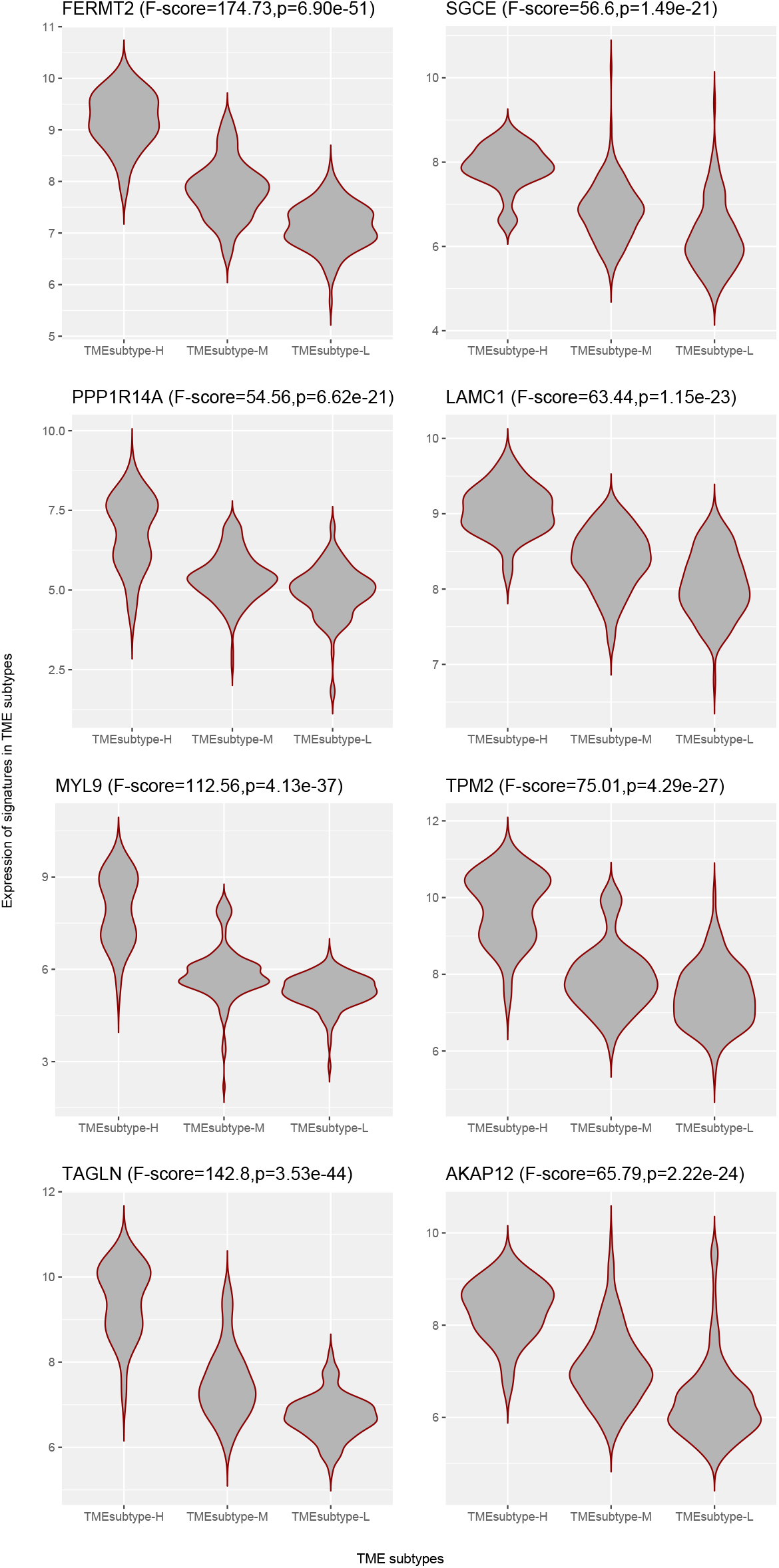
Violin plots showing differential expression of representative stromal cell markers in three TME subtypes.

The EMEC subtype showed an opposite trend. A lower abundance of that subtype has been shown to be associated with poorer prognosis. We therefore identified the prognostic genes for each GC cohort at a p-value cutoff of 0.05, as well as with a hazard ratio below 1, which suggests a significantly smaller risk. No significant prognostic genes were found across all four GC cohorts, therefore, we just kept the genes that were significantly prognostic across three of the four GC cohorts for further analysis. Finally, four significant prognostic gene signatures, including KCNQ1 (potassium voltage-gated channel subfamily Q member 1, HR=0.73), SURF6 (surfeit 6, HR=0.57), AGMAT (agmatinase, HR=0.79), and MRPS2 (mitochondrial ribosomal protein S2, HR=0.6), were detected after performing meta-analysis (*p* < 10^−5^, z-test for overall effect; Fig.21). Violin plots for the ACRG cohort indicated the representative prognostic marker genes were highly expressed in TMEsubtype-L with relatively low levels in other two TME subtypes (Fig.22). The expression levels were significantly different among three TME subtypes (*p* < 0.001, one-way ANOVA F test).

**Figure 21:**
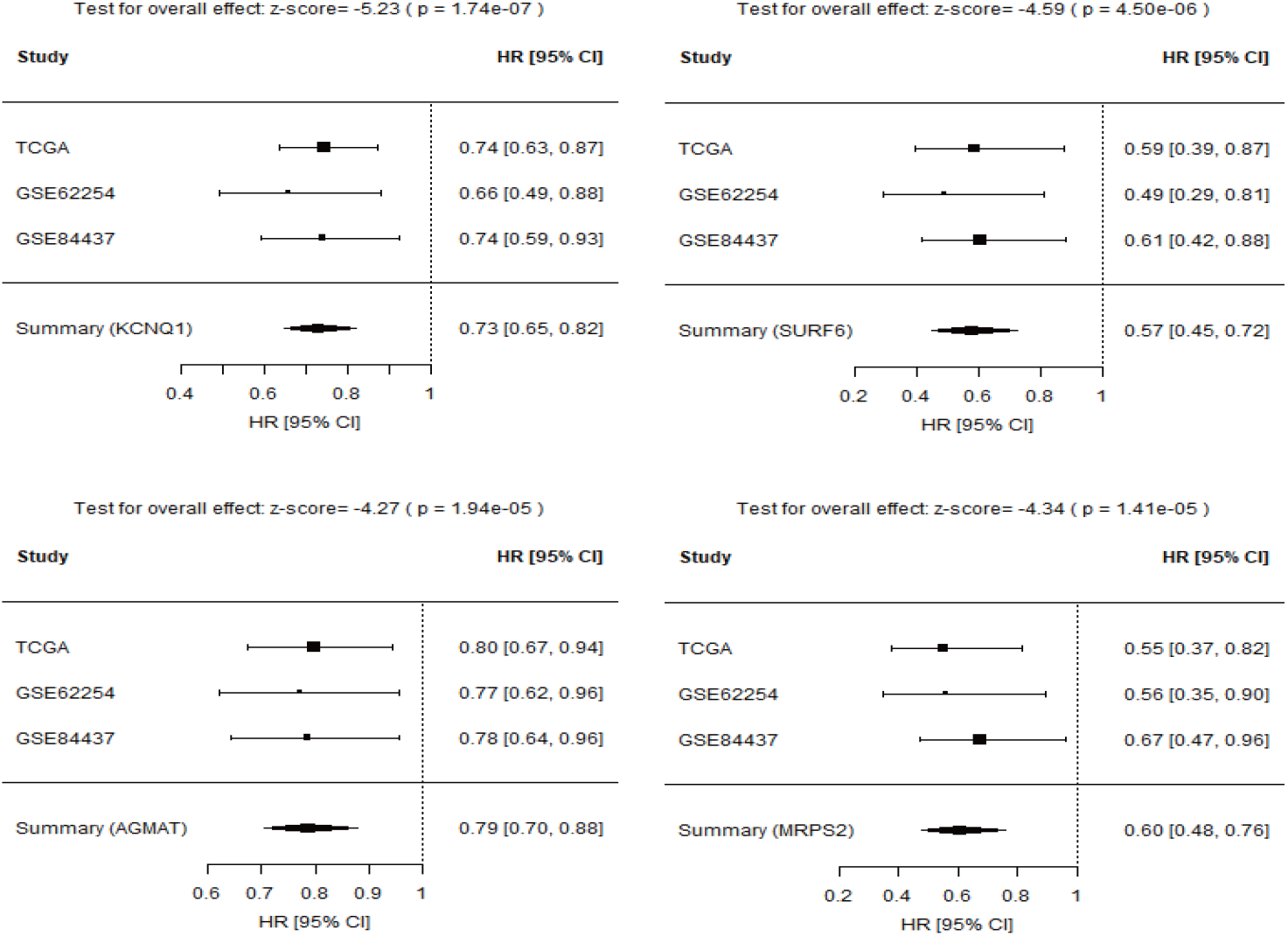
Forest plot of estimated hazard ratios of prognostic gene signatures in EMEC population.

**Figure 22:**
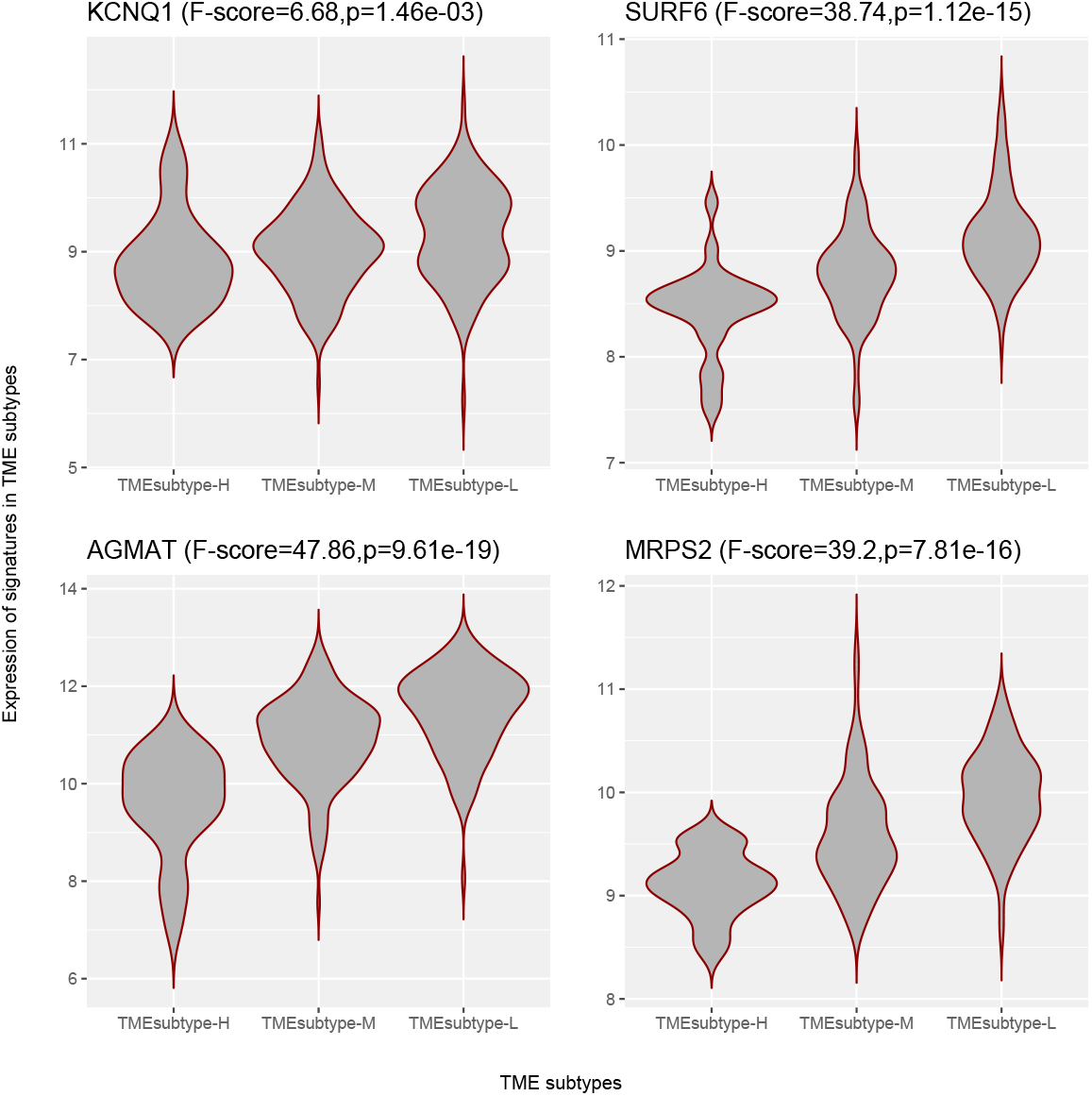
Violin plots showing differential expression of representative EMEC markers in three TME subtypes.

## 3 Discussion

In the present study, a comprehensive and systematic analysis of diverse epithelial, stromal, and immune cell types within the TME and their associations with GC risk was developed. We examined several large cohorts of GC patients at the cellular level and found a new and strong independent prognostic factor (STEM score) for GC patients. The STEM score was defined as the arithmetic sum of the two most significant TME factors: EMEC to stromal ratio and adaptive T cell to monocyte ratio. Our results suggest that high-risk patient groups (STEM score*≤* 3.95) have significantly shorter OS times than patients in the low-risk group (STEM score> 3.95).

Stromal cells, especially cancer-associated fibroblasts (CAFs), in the TME have been found to promote growth and survival of malignant cells [31]. Many studies have found that cancer cells release factors promoting fibroblasts to secrete tumor-promoting chemokines [32]. The interactions of tumors and CAFs can lead to increased malignancy in many cancer types [33,34]. Several studies of GC suggest that low tumor to stroma ratio (TSR) is associated with poor prognosis [19, 35]. Herein, we analyzed a gastric scRNA-seq data set that covered diverse epithelial cell types isolated from patients with NAG, CAG, IM, and EGC, and identified the EMEC population. The EMEC to stromal cell ratio was shown to have significant correlation to OS, which agreed with previous studies on the positive prognostic value of TSR. The EMEC population is comprised of cancer cells, MSCs, PCs, and PMCs that emerged uniformly in the EGC biopsy and were predominantly present in tumor samples but not typically found in adjacent normal samples. Additionally, significantly higher EMEC populations were detected in patients with stage I cancer than stage II, III, or IV, suggesting the value of the EMEC population in the early detection of gastric cancer.

There is increasing evidence that suggests a strong infiltration of T cells, especially CD8+ T cells, into the TME correlates with a good prognosis in many types of cancer and has implications for success of active cancer immunotherapy [36, 37]. Studies have shown that CD8+ T cells play a vital role in mediating anti-tumor immunity, and cytotoxic CD8+ memory T cells kill tumor cells by recognizing tumor-associated antigens presented on major histocompatibility complex class I [38–40]. High numbers of CD4+ T helper 1 cells in the TME also correlate with better prognosis [40]. Tumor-associated macrophages were found to enhance malignant cell migration, invasion, and metastases [41]. Monocytes can give rise to macrophages, so the abundance of monocytes may lead to increased production of macrophages. In our study of GC cohort, an increased adaptive T cell to monocyte ratio was significantly associated with increased OS. This is in line with studies by [42] of haematologic malignancies and [43] of stage III colon cancer. The studies of haematologic malignancies and stage III colon cancer demonstrate that an elevated lymphocyte to monocyte ratio (LMR) yields better survival outcome.

Molecular signatures associated with distinct clinical outcomes have been studied in many types of cancer [5,44,45]. We identified several gene signatures of the EMEC and stromal cell populations to be independent prognostic factors of OS in multivariate analysis. Several prognostic gene signatures of stromal cells have been previously reported to play oncogenic roles in cancer cell proliferation, migration, or invasion. FERMT2 (also known as Kindlin-2), a focal adhesion protein, has been found to regulate cancer cell proliferation, apoptosis, and chromosomal abnormalities in breast cancer that are associated with tumor stromal invasion, lymph node metastasis, and patient outcome in gastric cancer. Overexpression of FERMT2 promotes tumor formation in breast cancer and was linked with poorer patient outcomes [46, 47]. TAGLN is expressed in fibroblasts and smooth muscle, and the overexpression of TAGLN has been found in the tumor-induced reactive myofibroblastic stromal tissue in lung adenocarcinoma, as well in carcinomas of the stomach, liver, and oesophagus [48]. Silencing of TAGLN2, a homologue of TAGLN, has been reported to significantly inhibit cell proliferation and increase of apoptosis in bladder cancer [49]. MYL9 was previously found to be over-expressed in stages III and IV non-small cell lung cancer [50]. Overexpression of MYL9 in tumor cells was associated with poorer OS and recurrence-free survival in esophageal squamous cell carcinoma [51]. TPM2, a marker of fibroblast, was previously reported to be associated with poor prognosis in colorectal cancer [52]. The TAGLN, MYL9 and TPM2 were found to be over-expressed in fibroblasts from primary tumors compared to adjacent normal tissues and were associated with poorer prognosis in the TCGA cohort of colorectal cancer [52]. PPP1R14A has been investigated as a prognostic biomarker of gastric cancer [53]. LAMC1 was found to be a target of miR-29s. Silencing of LAMC1 significantly inhibited cell migration and invasion in prostate cancer cells [54]. AKAP12 has been investigated as a tumor suppressant in some human primary cancers, including GC [55,56], however, in the present study we found it significantly over-expressed in the TMEsubtype-H high risk group, suggesting an increased risk on OS with higher expression level.

Herein, we identified four prognostic gene signatures of EMEC population to be positively associated with OS. AGMAT were found to be positively associated with OS in kidney renal clear cell carcinoma [57]. MRPS2 encoding the mitochondrial ribosomal protein S2 was important for mitoribosome formation and stability, and mitochondrial translation. It was reported to predict poor OS in ovarian cancer patients [58]. However, we found that it was associated with better clinical outcome in GC patients. KCNQ1 has been shown to distribute widely and be functionally relevant in a variety of epithelial tissues [59]. There is preliminary evidence to suggest that KCNQ1 is a tumor suppressor in the stomach and colon [60, 61]. Low or loss expression of KCNQ1 was previously found to associated with poor disease-free survival in stage II and III colon cancer patients [62].

In conclusion, we identified significant prognostic factors and gene signatures among diverse epithelial, stromal, and immune cell populations in the GC TME. This study demonstrated the STEM score as a TME prognostic factor for GC. It showed that a lower STEM score was significantly associated with a shorter survival time. The entire GC cohort was stratified into three risk groups (high-, moderate-, and low-risk) based on the four STEM populations, which yielded incremental survival times. The risk stratification model may aid stratification of patients with stage III gastric cancer for radiation therapy.

## 4 Materials and Methods

### 4.1 Gastric cancer bulk gene expression data

We collected four GC gene expression datasets with the associated clinical, pathological, and outcome data: GSE62254 (ACRG), GSE15459, GSE84437, and TCGA-STAD (stomach adenocarcinoma). The ACRG and GSE15459 data sets contained gene expression profiles of 300 and 192 patients, respectively. The raw data (CEL files) of these two data sets were downloaded from Gene Expression Omnibus (GEO, www.ncbi.nlm.nih.gov/geo/). The CEL files were MAS5 normalized in the R environment using the affy software package. Both data sets were converted to gene-specific expression matrices using the R package hgu133plus2.db. For the GSE84437 data set, we directly downloaded its expression matrix after using quantile normalization from GEO. The R package illuminaHumanv3.db was used to translate probe identifications (IDs) to gene symbols. When multiple probes were present for one gene, we selected the probe with the highest average expression across the samples. The TCGA-STAD data set from The Cancer Genome Atlas (TCGA) was downloaded by using the getTCGA function of R package TCGA2STAT. This returned a gene expression matrix of RSEM [63] values.

The corresponding clinical, pathological, and outcome data of these data sets were collected as follows. We collected clinical data of the ACRG cohort from the supplementary materials of the original publication [4]. For the GSE15459 and GSE84437 cohorts clinical data was retrieved from the GEO database. We used the getTCGA function of R package TCGA2STAT to obtain clinical and OS data for the TCGA-STAD cohort.

### 4.2 Identification of ten non-immune cell populations from single-cell RNA-Seq of gastric antral mucosa biopsies

The single-cell RNA-seq data of patients with gastric premalignant lesions and early GC were downloaded from the GEO database with accession number GSE134520. This data set, profiled by 10X Chromium v2 (3’ assay), consists of 32,332 cells from nine patients with non-atrophic gastritis (NAG), chronic atrophic gastritis (CAG), intestinal metaplasia (IM), and early gastric cancer (EGC). We only included non-immune cell types with more than 20 cells’ sequencing data available at two or more stages for further analysis. During quality control, we further excluded cells with fewer than 500 expressed genes and removed genes detected in less than 2% cells, leaving 9926 genes in a total of 24,874 single cells.

To explore the hierarchical relationship among cell types in the cascade from gastritis to EGC, agglomerative hierarchical clustering was performed on the gastric scRNA-seq data set. The counts of single cell gene expression data were summarized across all cells mapped to the same cell type and subject. We then normalized the count summarization matrix by the transcripts per million (TPM) method. Next, we used the two-way analysis of variance (ANOVA) *F* test to identify cell type specific expressed genes. The main effects of cell type and subject were analyzed. The ANOVA analysis tested for differentially expressed genes between a cell type and all other cells. We selected the top 100 genes with high *F* value of cell type compared to *F* value of subject. We next computed the mean expression for the cell type specific expressed genes across expression profiles mapped to the same cell type and stage. The hierarchical cluster tree was generated using Pearson correlation coefficient 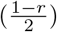 as the pairwise distance on the log-transformed mean expression profiles and Ward’s linkage distance as the cluster distance. Ten non-immune cell populations consisting of EMEC, PMEC, enteroendocrine, GMC, goblet cell, MSC, PC, PMC, endothelial cell, and stromal cell were identified from the cluster tree (Fig.1). The mapping of 11 non-immune cell types of patients with NAG, CAG, IM, and EGC to 10 cell populations is provided in Table 5 and Fig. 23.

**Table 4:**
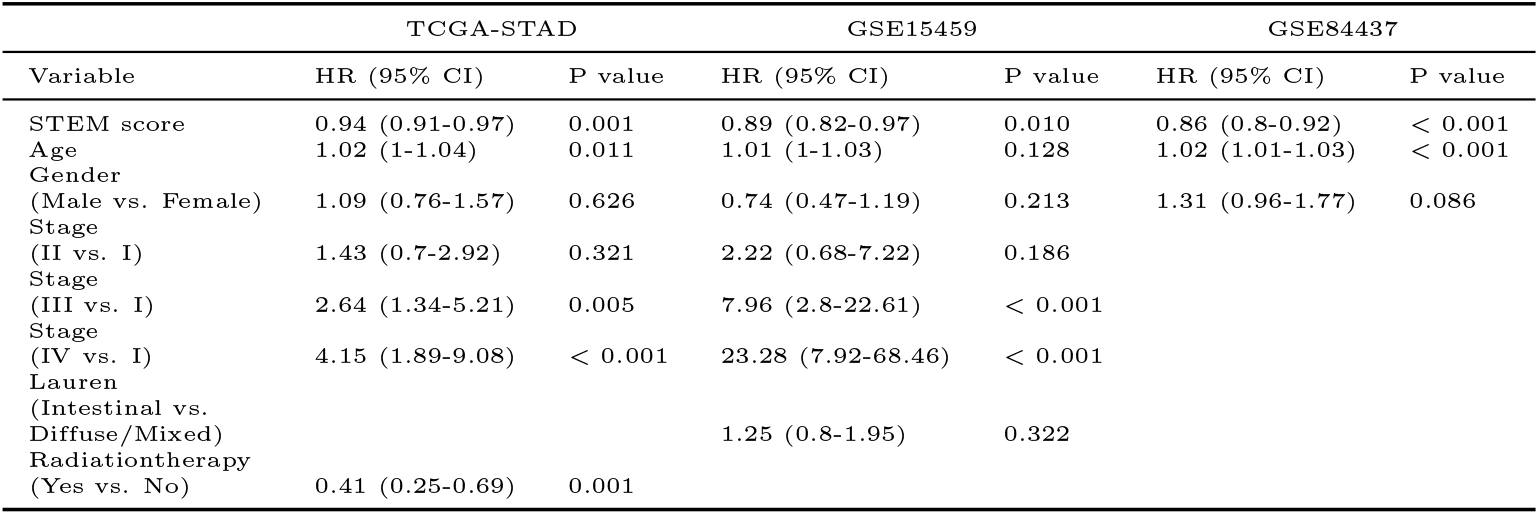
Multivariate Cox regression analysis used to examine the independent risk factor of prognosis in three independent validation cohorts.

**Table 5:** Grouping of the non-immune cell types for bulk gene expression samples deconvolution.

| Non-immune cell types | Full name | Stages | Grouping for Bulk deconvolution |
| --- | --- | --- | --- |
| Cancer cell | Cancer cell | EGC | EMEC |
| MSC | Metaplastic stem-like cell | EGC | EMEC |
| PC | Proliferative cell | EGC | EMEC |
| PMC | Pit mucous cell | EGC | EMEC |
| EC | Endothelial cell | CAG | Endothelial cell |
| EC | Endothelial cell | EGC | Endothelial cell |
| EC | Endothelial cell | IM | Endothelial cell |
| EC | Endothelial cell | NAG | Endothelial cell |
| Enteroendocrine | Enteroendocrine cell | CAG | Enteroendocrine |
| Enteroendocrine | Enteroendocrine cell | EGC | Enteroendocrine |
| Enteroendocrine | Enteroendocrine cell | IM | Enteroendocrine |
| Enteroendocrine | Enteroendocrine cell | NAG | Enteroendocrine |
| GMC | Antral basal gland mucous cell | CAG | GMC |
| GMC | Antral basal gland mucous cell | EGC | GMC |
| GMC | Antral basal gland mucous cell | IM | GMC |
| GMC | Antral basal gland mucous cell | NAG | GMC |
| Goblet cell | Goblet cell | CAG | Goblet cell |
| Goblet cell | Goblet cell | EGC | Goblet cell |
| Goblet cell | Goblet cell | IM | Goblet cell |
| Goblet cell | Goblet cell | NAG | Goblet cell |
| MSC | Metaplastic stem-like cell | CAG | MSC |
| MSC | Metaplastic stem-like cell | NAG | MSC |
| PC | Proliferative cell | CAG | PC |
| PC | Proliferative cell | IM | PC |
| PC | Proliferative cell | NAG | PC |
| PMC | Pit mucous cell | CAG | PMC |
| PMC | Pit mucous cell | IM | PMC |
| PMC | Pit mucous cell | NAG | PMC |
| Cancer cell | Cancer cell | CAG | PMEC |
| Cancer cell | Cancer cell | IM | PMEC |
| Enterocyte | Enterocyte | CAG | PMEC |
| Enterocyte | Enterocyte | EGC | PMEC |
| Enterocyte | Enterocyte | IM | PMEC |
| Enterocyte | Enterocyte | NAG | PMEC |
| Fibroblast | Fibroblast | CAG | Stromal cell |
| Fibroblast | Fibroblast | EGC | Stromal cell |
| Fibroblast | Fibroblast | IM | Stromal cell |
| Fibroblast | Fibroblast | NAG | Stromal cell |
| SM cell | Smooth muscle cell | CAG | Stromal cell |
| SM cell | Smooth muscle cell | IM | Stromal cell |
| SM cell | Smooth muscle cell | NAG | Stromal cell |
| Cancer cell | Cancer cell | NAG | Undefined |
| MSC | Metaplastic stem-like cell | IM | Undefined |

**Figure 23:**
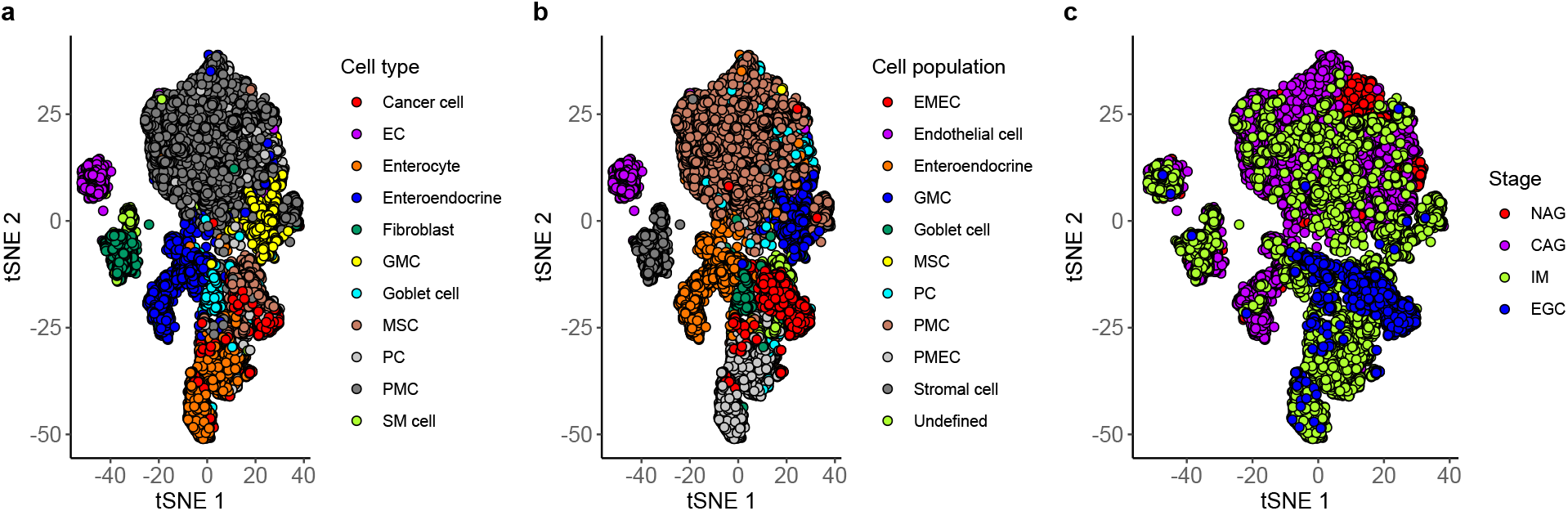
The t-SNE plot of whole transcriptomes of 24874 cells from gastric antral mucosa biopsies. (a) The 11 cell types are denoted by distinct colours. (b) The 10 identified cell populations are denoted by distinct colours. (c) The 4 stages are denoted by distinct colours.

### 4.3 Marker selection and signature matrix construction

To estimate the proportions of non-immune cell populations in the gastric samples we created a signature matrix composed of the characteristic expression profiles for each of the 10 non-immune cell populations. This signature matrix distinguished EMEC, PMEC, enteroendocrine, GMC, goblet cell, MSC, PC, PMC, endothelial cell, and stromal cell populations. The matrix was generated based on the gastric single cell data. The counts of single cell gene expression data were summarized across all cells mapped to the same cell type and subject, then normalized to the count summarization matrix by the TPM method. The expression profiles were averaged within each cell population. This generated a matrix of genes × cell populations. The signature matrix was defined as the sub-matrix formed by a set of cell population-specific marker genes. The marker genes were selected by first selecting genes with a two-fold or higher over differential expressed between one cell population and all other cell populations, then filtering out non-significant genes with a p-value larger than 0.01 (one-way ANOVA analysis tested for each gene between a cell population and all other cells). Next, we ranked genes in decreasing order by their fold changes and selected the top 150 genes for each cell population, which resulted in a signature matrix of 1319 genes by 10 cell populations. The signature matrix and gene signatures for each non-immune cell population are available on Github (https://github.com/wenjshen/STEM).

To quantify the proportions of immune cell populations, we used a signature matrix provided in [64] consisting of 1296 genes in 17 immune cell types whose transcriptomic profiling was sorted by RNA-seq. This signature matrix covers the majority of cells that constitute a PBMC sample. We then merged these 17 immune cell types into 7 major lineages according to their biological similarity, resulting in 7 immune cell populations: adaptive T cells, innate T cells, adaptive B cells, natural killer cells, monocytes, dendritic cells, and granulocytes (Table 6).

**Table 6:** Grouping of the immune cell types for bulk gene expression samples deconvolution.

| Immune cell types | Full name | Grouping for Bulk deconvolution |
| --- | --- | --- |
| Monocytes C | Classical monocytes | Monocytes |
| Monocytes NC+I | Non-classical/intermediate monocytes | Monocytes |
| NK | Natural killer cells | NK |
| T gd non-Vd2 | $\gamma\delta$ non-Vδ2 T cells | T Innate |
| T gd Vd2 | $\gamma\delta$ Vδ2 T cells | T Innate |
| Neutrophils LD | Low-density neutrophils | Granulocytes |
| Basophils LD | Low-density basophils | Granulocytes |
| mDCs | Myeloid dendritic cells | DCs |
| pDCs | Plasmacytoid dendritic cells | DCs |
| T CD8 Naive | Nave CD8 T cells | T adaptive |
| T CD8 Memory | Memory CD8 T cells | T adaptive |
| B Naive | Naive B cells | B adaptive |
| B Memory | Memory B cells | B adaptive |
| Plasmablasts | Plasmablasts | B adaptive |
| T CD4 Naive | Nave CD4 T cells | T adaptive |
| T CD4 Memory | Memory CD4 T cells | T adaptive |
| MAIT | Mucosal associated invariant T cells | T Innate |

### 4.4 Deconvolving bulk gene expression samples

For the deconvolution of the bulk RNA-seq data, the expression profiles of bulk samples were normalized for sequencing depth and gene length using the Transcripts Per Million (TPM) method [65]. We performed deconvolution with support vector regression using the CIBERSORT algorithm [25].

The microarray data was quantile normalized. We next used the CIBERSORTx method [23] to deconvolve bulk samples. Additionally, batch correction was applied to reduce cross-platform variance.

The deconvolution method was combined with the signature matrices of non-immune and immune cell populations, respectively, to estimate non-immune and immune cell relative fractions of GC samples. Since CIBERSORT and CIBERSORTx estimated relative but not absolute fractions of cell populations within a sample, the values may not be comparable across samples. We therefore defined non-immune scores (or immune scores) by calculating the ratios of relative fractions in each pair of non-immune (or immune) cell populations. The non-immune or immune scores are therefore comparable across samples.

### 4.5 GO enrichment analyses

The functional classification of the EMEC gene signatures was analyzed in the GO. The R package clusterProfiler [66] (version 3.18.1) was used to identify and visualize enriched GO terms. Significance in all enrichment analyses were based on BH-corrected *p* value < 0.1 and the gene counts ≥ 2.

### 4.6 Determination of Optimal STEM score cutoff

To identify the statistically optimal cutoff of the STEM score, we analyzed the influence of the STEM score on survival outcome in the ACRG cohort by using univariate Cox proportional hazard modeling. The analysis was performed for a dense set of quantiles from 0.1th to 0.9th, with a 0.01 step. Each analysis divided the entire cohort into two groups, and the cutoff that minimized the P-value for testing the risk difference between the two groups was selected.

### 4.7 TME subtypes identification in GC

Each GC sample is represented by four input features of the estimated relative abundance of the cell populations: EMECs, stromal cells, adaptive T cells, and monocytes. Spectral clustering was performed on the meta-cohort (ACRG, GSE15459, GSE84437, and TCGA-STAD) by using the radial basis kernel function to measure the similarity between two samples, which was implemented in the R kernlab package. The optimal number of clusters was chosen using the NbClust function, which was implemented in the R NbClust [67] package. NbClust utilizes 26 different cluster validity indices with Euclidean as the distance measurement method and Ward’s hierarchical clustering as the clustering method to generate a majority rules number of clusters for the GC data set.

### 4.8 Statistical analysis

All the statistical analyses were performed in the R environment (version 4.0.3). Cumulative survival time was calculated using the Kaplan-Meier method and the differences in survival curves were analyzed using the log-rank test from R package survminer. Univariate and multivariate analyses were conducted using the Cox proportional hazards regression modeling using the R package survival. For all tests, the *p*-value cutoff for statistical significance was set as 0.05 as default unless an alternative value is specified. Statistical significance between tumor samples and adjacent normal samples was assessed using Student’s t test and indicated as follows: ^*\**^*p* < 0.05,^*\*\**^ *p* < 0.01,^*\*\*\**^ *p* < 0.001,^*\*\*\*\**^ *p* < 0.0001.

## Data Availability

The data analyzed in this study are available from the Gene Expression Omnibus (accession numbers: GSE62254; GSE15459; GSE84437; GSE134520), The Cancer Genome Atlas Project or from the authors upon reasonable request.

## Acknowledgments

This work is supported by Vanderbilt University Development Funds (FF 300033).

## References

[1] Jacques Ferlay, M Colombet, I Soerjomataram, C Mathers, DM Parkin,Piñeros, A Znaor, and F Bray. Estimating the global cancer incidence and mortality in 2018: Globocan sources and methods. International journal of cancer, 144(8):1941–1953, 2019.

[2] Pekka Lauren. The two histological main types of gastric carcinoma: diffuse and so-called intestinal-type carcinoma: an attempt at a histo-clinical classification. Acta Pathologica Microbiologica Scandinavica, 64(1):31–49, 1965.

[3] Ming Teh and Yoke-Sun Lee. Intestinal and diffuse carcinoma of the stomach among the ethnic and dialect groups in singapore. Cancer, 60(4):921–925, 1987.

[4] Razvan Cristescu, Jeeyun Lee, Michael Nebozhyn, Kyoung-Mee Kim, Jason C Ting, Swee Seong Wong, Jiangang Liu, Yong Gang Yue, Jian Wang, Kun Yu, et al. Molecular analysis of gastric cancer identifies subtypes associated with distinct clinical outcomes. Nature medicine, 21(5):449–456, 2015.

[5] Cancer Genome Atlas Research Network et al. Comprehensive molecular characterization of gastric adenocarcinoma. Nature, 513(7517):202–209, 2014.

[6] Gloria H Heppner and Bonnie E Miller. Tumor heterogeneity: biological implications and therapeutic consequences. Cancer and Metastasis Reviews, 2(1):5–23, 1983.

[7] Ash A Alizadeh, Victoria Aranda, Alberto Bardelli, Cedric Blanpain, Christoph Bock, Christine Borowski, Carlos Caldas, Andrea Califano, Michael Doherty, Markus Elsner, et al. Toward understanding and exploiting tumor heterogeneity. Nature medicine, 21(8):846, 2015.

[8] Shannon J Turley, Viviana Cremasco, and Jillian L Astarita. Immunological hallmarks of stromal cells in the tumour microenvironment. Nature reviews immunology, 15(11):669–682, 2015.

[9] Raghu Kalluri. The biology and function of fibroblasts in cancer. Nature Reviews Cancer, 16(9):582, 2016.

[10] Wolf H Fridman, Laurence Zitvogel, Catherine Sautes-Fridman, and Guido Kroemer. The immune contexture in cancer prognosis and treatment. Nature reviews Clinical oncology, 14(12):717, 2017.

[11] Tianyi Liu, Linli Zhou, Danni Li, Thomas Andl, and Yuhang Zhang. Cancer-associated fibroblasts build and secure the tumor microenvironment. Frontiers in cell and developmental biology, 7:60, 2019.

[12] TL Whiteside. The tumor microenvironment and its role in promoting tumor growth. Oncogene, 27(45):5904–5912, 2008.

[13] Frances R Balkwill, Melania Capasso, and Thorsten Hagemann. The tumor microenvironment at a glance, 2012.

[14] Yu Yan, Ruifen Wang, Wenbin Guan, Meng Qiao, and Lifeng Wang. Roles of micrornas in cancer associated fibroblasts of gastric cancer. Pathology-Research and Practice, 213(7):730–736, 2017.

[15] Jing Zhai, Jiajia Shen, Guiping Xie, Jiaqi Wu, Mingfang He, Lili Gao, Yifen Zhang, Xuequan Yao, and Lizong Shen. Cancer-associated fibroblasts-derived il-8 mediates resistance to cisplatin in human gastric cancer. Cancer letters, 454:37–43, 2019.

[16] Dachuan Zhang, Wenting He, Chao Wu, Yan Tan, Yang He, Bin Xu, Lujun Chen, Qing Li, and Jingting Jiang. Scoring system for tumor-infiltrating lymphocytes and its prognostic value for gastric cancer. Frontiers in immunology, 10:71, 2019.

[17] Zongxin Ling, Li Shao, Xia Liu, Yiwen Cheng, Chongxian Yan, Ying Mei, Feng Ji, and Xiaosun Liu. Regulatory t cells and plasmacytoid dendritic cells within the tumor microenvironment in gastric cancer are correlated with gastric microbiota dysbiosis: a preliminary study. Frontiers in immunology, 10:533, 2019.

[18] Peiming Zheng, Qin Luo, Weiwei Wang, Junhua Li, Tingting Wang, Ping Wang, Lei Chen, Peng Zhang, Hui Chen, Yi Liu, et al. Tumor-associated macrophages-derived exosomes promote the migration of gastric cancer cells by transfer of functional apolipoprotein e. Cell death & disease, 9(4):1–14, 2018.

[19] Chunwei Peng, Jiuyang Liu, Guifang Yang, and Yan Li. The tumor-stromal ratio as a strong prognosticator for advanced gastric cancer patients: proposal of a new tsnm staging system. Journal of gastroenterology, 53(5):606–617, 2018.

[20] Giuseppe Sammarco, Gilda Varricchi, Valentina Ferraro, Michele Ammendola, Michele De Fazio, Donato Francesco Altomare, Maria Luposella, Lorenza Maltese, Giuseppe Currò, Gianni Marone, et al. Mast cells, angiogenesis and lymphangiogenesis in human gastric cancer. International journal of molecular sciences, 20(9):2106, 2019.

[21] Bailiang Li, Yuming Jiang, Guoxin Li, George A Fisher Jr, and Ruijiang Li. Natural killer cell and stroma abundance are independently prognostic and predict gastric cancer chemotherapy benefit. JCI insight, 5(9), 2020.

[22] Xuran Wang, Jihwan Park, Katalin Susztak, Nancy R Zhang, and Mingyao Li. Bulk tissue cell type deconvolution with multi-subject single-cell expression reference. Nature communications, 10(1):1–9, 2019.

[23] Aaron M Newman, Chloé B Steen, Chih Long Liu, Andrew J Gentles, Aadel A Chaudhuri, Florian Scherer, Michael S Khodadoust, Mohammad S Esfahani, Bogdan A Luca, David Steiner, et al. Determining cell type abundance and expression from bulk tissues with digital cytometry. Nature biotechnology, 37(7):773–782, 2019.

[24] Peng Zhang, Mingran Yang, Yiding Zhang, Shuai Xiao, Xinxing Lai, Aidi Tan, Shiyu Du, and Shao Li. Dissecting the single-cell transcriptome network underlying gastric premalignant lesions and early gastric cancer. Cell reports, 27(6):1934–1947, 2019.

[25] Aaron M Newman, Chih Long Liu, Michael R Green, Andrew J Gentles, Weiguo Feng, Yue Xu, Chuong D Hoang, Maximilian Diehn, and Ash A Alizadeh. Robust enumeration of cell subsets from tissue expression profiles. Nature methods, 12(5):453–457, 2015.

[26] John P Jakupciak, Samantha Maragh, Maura E Markowitz, Alissa K Greenberg, Mohammad O Hoque, Anirban Maitra, Peter E Barker, Paul D Wagner, William N Rom, Sudhir Srivastava, et al. Performance of mitochondrial dna mutations detecting early stage cancer. BMC cancer, 8(1):1–11, 2008.

[27] Meghan L Verschoor, Robert Ungard, Andrew Harbottle, John P Jakupciak, RL Parr, and Gurmit Singh. Mitochondria and cancer: past, present, and future. BioMed research international, 2013, 2013.

[28] Hang Yang, Yan Li, and Bing Hu. Potential role of mitochondria in gastric cancer detection: Fission and glycolysis. Oncology Letters, 21(6):1–7, 2021.

[29] M Constanza Camargo, Woo-Ho Kim, Anna Maria Chiaravalli, Kyoung-Mee Kim, Alejandro H Corvalan, Keitaro Matsuo, Jun Yu, Joseph JY Sung, Roberto Herrera-Goepfert, Fernando Meneses-Gonzalez, et al. Improved survival of gastric cancer with tumour epstein–barr virus positivity: an international pooled analysis. Gut, 63(2):236–243, 2014.

[30] LIN Zhu, ZHI Li, YAN Wang, Chenlu Zhang, Yunpeng Liu, and Xiujuan Qu. Microsatellite instability and survival in gastric cancer: A systematic review and meta-analysis. Molecular and clinical oncology, 3(3):699–705, 2015.

[31] Daniela F Quail and Johanna A Joyce. Microenvironmental regulation of tumor progression and metastasis. Nature medicine, 19(11):1423–1437, 2013.

[32] Pravin Mishra, Debabrata Banerjee, and Adit Ben-Baruch. Chemokines at the crossroads of tumor-fibroblast interactions that promote malignancy. Journal of leukocyte biology, 89(1):31–39, 2011.

[33] Akira Orimo, Piyush B Gupta, Dennis C Sgroi, Fernando Arenzana-Seisdedos, Thierry Delaunay, Rizwan Naeem, Vincent J Carey, Andrea L Richardson, and Robert A Weinberg. Stromal fibroblasts present in invasive human breast carcinomas promote tumor growth and angiogenesis through elevated sdf-1/cxcl12 secretion. Cell, 121(3):335–348, 2005.

[34] Laurie E Littlepage, Mikala Egeblad, and Zena Werb. Coevolution of cancer and stromal cellular responses. Cancer cell, 7(6):499–500, 2005.

[35] Niko Kemi, Maarit Eskuri, Anni Herva, Joni Leppänen, Heikki Huhta, Olli Helminen, Juha Saarnio, Tuomo J Karttunen, and Joonas H Kauppila. Tumour-stroma ratio and prognosis in gastric adenocarcinoma. British journal of cancer, 119(4):435–439, 2018.

[36] Sahar MA Mahmoud, Emma Claire Paish, Desmond G Powe, R Douglas Macmillan, Matthew J Grainge, Andrew HS Lee, Ian O Ellis, and Andrew R Green. Tumor-infiltrating cd8+ lymphocytes predict clinical outcome in breast cancer. Journal of clinical oncology, 29(15):1949–1955, 2011.

[37] John Reiser and Arnob Banerjee. Effector, memory, and dysfunctional cd8+ t cell fates in the antitumor immune response. Journal of immunology research, 2016, 2016.

[38] Steven A Rosenberg. Progress in human tumour immunology and immunotherapy. Nature, 411(6835):380–384, 2001.

[39] TA Zikos, AD Donnenberg, RJ Landreneau, JD Luketich, and VS Donnenberg. Lung t-cell subset composition at the time of surgical resection is a prognostic indicator in non-small cell lung cancer. Cancer immunology, immunotherapy, 60(6):819–827, 2011.

[40] Wolf Herman Fridman, Franck Pagès, Catherine Sautès-Fridman, and Jérôme Galon. The immune contexture in human tumours: impact on clinical outcome. Nature Reviews Cancer, 12(4):298–306, 2012.

[41] John Condeelis and Jeffrey W Pollard. Macrophages: obligate partners for tumor cell migration, invasion, and metastasis. Cell, 124(2):263–266, 2006.

[42] Zhi-Ming Li, Jia-Jia Huang, Yi Xia, Jian Sun, Ying Huang, Yu Wang, Ying-Jie Zhu, Ya-Jun Li, Wei Zhao, Wen-Xiao Wei, et al. Blood lymphocyte-to-monocyte ratio identifies high-risk patients in diffuse large b-cell lymphoma treated with r-chop. PloS one, 7(7):e41658, 2012.

[43] M Stotz, M Pichler, G Absenger, J Szkandera, F Arminger, R Schaberl-Moser, H Samonigg, T Stojakovic, and A Gerger. The preoperative lymphocyte to monocyte ratio predicts clinical outcome in patients with stage iii colon cancer. British journal of cancer, 110(2):435–440, 2014.

[44] Dong Keun Rhee, Su Hyung Park, and Yeun Kyu Jang. Molecular signatures associated with transformation and progression to breast cancer in the isogenic mcf10 model. Genomics, 92(6):419–428, 2008.

[45] Timothy R Donahue, Linh M Tran, Reginald Hill, Yunfeng Li, Anne Kovochich, Joseph H Calvopina, Sanjeet G Patel, Nanping Wu, Antreas Hindoyan, James J Farrell, et al. Integrative survival-based molecular profiling of human pancreatic cancer. Clinical Cancer Research, 18(5):1352–1363, 2012.

[46] Zhanlong Shen, Yingjiang Ye, Lingyi Dong, Sanna Vainionpää, Harri Mustonen, Pauli Puolakkainen, and Shan Wang. Kindlin-2: a novel adhesion protein related to tumor invasion, lymph node metastasis, and patient outcome in gastric cancer. The American journal of surgery, 203(2):222–229, 2012.

[47] Ting Zhao, Lizhao Guan, Yu Yu, Xuelian Pei, Jun Zhan, Ling Han, Yan Tang, Feng Li, Weigang Fang, and Hongquan Zhang. Kindlin-2 promotes genome instability in breast cancer cells. Cancer letters, 330(2):208–216, 2013.

[48] Jung-hyun Rho, Michael HA Roehrl, and Julia Y Wang. Tissue proteomics reveals differential and compartment-specific expression of the homologs transgelin and transgelin-2 in lung adenocarcinoma and its stroma. Journal of proteome research, 8(12):5610–5618, 2009.

[49] H Yoshino, T Chiyomaru, H Enokida, K Kawakami, S Tatarano, K Nishiyama, N Nohata, N Seki, and M Nakagawa. The tumour-suppressive function of mir-1 and mir-133a targeting tagln2 in bladder cancer. British journal of cancer, 104(5):808–818, 2011.

[50] Xiang Tan and Mingwu Chen. Mylk and myl9 expression in non-small cell lung cancer identified by bioinformatics analysis of public expression data. Tumor Biology, 35(12):12189–12200, 2014.

[51] Jian-Hua Wang, Lan Zhang, Shu-Ting Huang, Jing Xu, Yun Zhou, Xing-Juan Yu, Rong-Zhen Luo, Zhe-Sheng Wen, Wei-Hua Jia, and Min Zheng. Expression and prognostic significance of myl9 in esophageal squamous cell carcinoma. PLoS One, 12(4):e0175280, 2017.

[52] Yuan Zhou, Shuhui Bian, Xin Zhou, Yueli Cui, Wendong Wang, Lu Wen, Limei Guo, Wei Fu, and Fuchou Tang. Single-cell multiomics sequencing reveals prevalent genomic alterations in tumor stromal cells of human colorectal cancer. Cancer Cell, 38(6):818–828, 2020.

[53] Jun-Yi Hou, Yu-Gang Wang, Shi-Jie Ma, Bing-Yin Yang, and Qian-Ping Li. Identification of a prognostic 5-gene expression signature for gastric cancer. Journal of cancer research and clinical oncology, 143(4):619–629, 2017.

[54] Rika Nishikawa, Yusuke Goto, Satoko Kojima, Hideki Enokida, Takeshi Chiyomaru, Takashi Kinoshita, Shinichi Sakamoto, Miki Fuse, Masayuki Nakagawa, Yukio Naya, et al. Tumor-suppressive microrna-29s inhibit cancer cell migration and invasion via targeting lamc1 in prostate cancer. International journal of oncology, 45(1):401–410, 2014.

[55] Moon-Chang Choi, Hyun-Soon Jong, Tai Young Kim, Sang-Hyun Song, Dong Soon Lee, Jung Weon Lee, Tae-You Kim, Noe Kyeong Kim, and Yung-Jue Bang. Akap12/gravin is inactivated by epigenetic mechanism in human gastric carcinoma and shows growth suppressor activity. Oncogene, 23(42):7095–7103, 2004.

[56] Irwin H Gelman. Emerging roles for ssecks/gravin/akap12 in the control of cell proliferation, cancer malignancy, and barriergenesis. Genes & cancer, 1(11):1147–1156, 2010.

[57] Xudong Guo, Zhuolun Sun, Shaobo Jiang, Xunbo Jin, and Hanbo Wang. Identification and validation of a two-gene metabolic signature for survival prediction in patients with kidney renal clear cell carcinoma. Aging (Albany NY), 13(6):8276, 2021.

[58] Federica Sotgia and Michael P Lisanti. Mitochondrial mrna transcripts predict overall survival, tumor recurrence and progression in serous ovarian cancer: companion diagnostics for cancer therapy. Oncotarget, 8(40):66925, 2017.

[59] Markus Bleich and Richard Warth. The very small-conductance k+ channel k v lqt1 and epithelial function. Pflügers Archiv, 440(2):202–206, 2000.

[60] Bich LN Than, JACM Goos, Aaron L Sarver, Michael Gerard O’Sullivan, Annette Rod, Timothy Kaehler Starr, Remond JA Fijneman, Gerrit A Meijer, Lei Zhao, Yuanyuan Zhang, et al. The role of kcnq1 in mouse and human gastrointestinal cancers. Oncogene, 33(29):3861–3868, 2014.

[61] Raphael Rapetti-Mauss, Viviana Bustos, Warren Thomas, Jean McBryan, Harry Harvey, Natalia Lajczak, Stephen F Madden, Bernard Pellissier, Franck Borgese, Olivier Soriani, et al. Bidirectional kcnq1: β-catenin interaction drives colorectal cancer cell differentiation. Proceedings of the National Academy of Sciences, 114(16):4159–4164, 2017.

[62] Sjoerd H Den Uil, Veerle MH Coupé, Janneke F Linnekamp, Evert Van Den Broek, Jeroen ACM Goos, Pien M Delis-van Diemen, J Eric, Nicole CT Van Grieken, Patricia M Scott, Louis Vermeulen, et al. Loss of kcnq1 expression in stage ii and stage iii colon cancer is a strong prognostic factor for disease recurrence. British journal of cancer, 115(12):1565–1574, 2016.

[63] Bo Li and Colin N Dewey. Rsem: accurate transcript quantification from rna-seq data with or without a reference genome. BMC bioinformatics, 12(1):323, 2011.

[64] Gianni Monaco, Bernett Lee, Weili Xu, Seri Mustafah, You Yi Hwang, Christophe Carre, Nicolas Burdin, Lucian Visan, Michele Ceccarelli, Michael Poidinger, et al. Rna-seq signatures normalized by mrna abundance allow absolute deconvolution of human immune cell types. Cell reports, 26(6):1627–1640, 2019.

[65] Bo Li, Victor Ruotti, Ron M Stewart, James A Thomson, and Colin N Dewey. Rna-seq gene expression estimation with read mapping uncertainty. Bioinformatics, 26(4):493–500, 2010.

[66] Guangchuang Yu, Li-Gen Wang, Yanyan Han, and Qing-Yu He. clusterprofiler: an r package for comparing biological themes among gene clusters. Omics: a journal of integrative biology, 16(5):284–287, 2012.

[67] Charrad Malika, Nadia Ghazzali, Veronique Boiteau, and Azam Niknafs. Nbclust: an r package for determining the relevant number of clusters in a data set. J. Stat. Softw, 61:1–36, 2014.

